# Identification of common variants influencing risk of the three-repeat tauopathy Pick’s disease: a genome wide association study

**DOI:** 10.64898/2025.12.21.25342758

**Authors:** William J Scotton, Rebecca R Valentino, Alejandro Martinez-Carrasco, Raquel Real, Hannah L Macpherson, Nicole Tamvaka, Kin Mok, Michael G Heckman, Christopher Kobylecki, Valentina Escott-Price, James B Rowe, Huw R Morris, Rosa Rademakers, Shanu F Roemer, Tammaryn Lashley, Dennis W Dickson, Jonathan D Rohrer, John A Hardy, Owen A Ross, Maryam Shoai, the Pick’s International Consortium (PIC)

## Abstract

Pick’s disease (PiD) is a rare cause of sporadic frontotemporal dementia, neuropathologically defined by the presence of Pick bodies consisting of aggregates of 3-repeat tau. Given the genetic aetiology of PiD remains unresolved, we assembled the Pick’s disease International Consortium (PIC) to identify susceptibility loci through a genome-wide association study (GWAS). A GWAS was conducted in 294 autopsy confirmed PiD cases and 1,055 controls. Lead variants were annotated using the Functional Mapping and Annotation of GWAS (FUMA) platform, followed by co-localisation analyses using the METABRAIN dataset and statistical finemapping using FINEMAP and SuSiE. After exclusion of 3 cases of MAPT mutations, no variants were associated with risk of PiD at genome-wide significance (*p* < 5 × 10^-8^). The strongest association was on chromosome 4 (4p13, lead SNP rs112161979, OR = 7.53, 95% Confidence Interval (CI) = 3.62-15.65, *p* = 6.37 × 10^-8^) followed by chromosome 11 (11p15.4, lead SNP rs66481907, OR = 2.10, 95% CI = 1.54-2.84, *p* = 1.83 × 10^-6^). rs112161979 is an intronic SNP in the *KCDT8* gene, encoding a potassium channel tetramerization domain that acts as an auxiliary subunit for GABA_B_ receptors, whilst rs66481907 is an intronic SNP in *TRIM22*, encoding an E3 ubiquitin-protein ligase. Our GWAS provides the first evidence of possible genetic risk for PiD that implicate the modulation of GABA_B_ receptor signalling and inflammation, in disease pathogenesis. Replication of these findings will be important, but our results suggest that, if present, the genetic risk of PiD beyond MAPT mutations is low.

## Introduction

Pick’s disease (PiD) is a rare cause of frontotemporal dementia. It is characterised by severe “knife-edge” frontal and temporal lobe atrophy and classified neuropathogically by the presence of ballooned neurons and argyrophilic inclusions called Pick bodies. These eponymous Pick bodies contain hyperphosphorylated 3-repeat tau aggregates, leading to its designation as a 3-repeat (3R) tauopathy, in contrast to the 4-repeat (4R) tauopathies such as progressive supranuclear palsy (PSP) and corticobasal degeneration (CBD). A recently proposed structure-based classification of the tauopathies, derived using cryo-electron microscopy, demonstrates that these 3R tau aggregates consist of a distinct disease-specific molecular conformation of tau fibrils in PiD(1,2).

Tau is encoded by the *MAPT* gene on chromosome 17, with six major isoforms in the adult human brain(3) generated through alternative splicing of exons 2, 3, and 10. Alternative splicing of exons 2 or 3 produces isoforms with either none, one or 2 amino terminal inserts, whereas alternative splicing of exon 10 produces isoforms with either 3R or 4R microtubule binding regions. Rare mutations in the *MAPT* gene can cause Pick’s-like 3R pathology(4,5), though there has been no systematic study of large cohorts to ascertain their true prevalence in PiD. The genomic architecture of the *MAPT* locus is characterised by two haplotypes resulting from a 900kb inversion of the H2 haplotype (6). Inheritance of the H1 haplotype has long been known to be a risk factor for both PSP (Odds Ratio [OR] = 5.46)(7,8) and CBD(9,10) (OR = 3.45), while more recently the H2 haplotype has been shown to be associated with an increased risk of Pick’s disease (OR 1.35)(11).

The rarity of PiD, combined with the difficulty of diagnosing the underlying pathology in life (due to lack of in vivo biomarkers and limited clinico-pathological correlations), has impeded large scale genetic studies in this disease(12,13). This is in contrast to the 4R tauopathies where there have been numerous genome wide association studies (GWAS), including three case-control studies (one in CBD and two in PSP)(8,10,14), an investigation of genetic determinants of PSP phenotype(15), and an evaluation of associations between genetic variation and survival in PSP(16). There have been no equivalent studies yet performed in PiD. The Pick’s International Consortium (PIC)(11) has collated the largest number of pathology-confirmed PiD cases to date, providing the opportunity to perform genome-wide association studies. Here we perform the first GWAS in PiD, using autopsy-confirmed cases from the PIC, and subsequently perform functional annotation and fine-mapping to better understand how the nominated genetic variants are associated with the regulation of gene expression and the underlying pathophysiology of disease.

## Materials and Methods

### Study population

321 neuropathogically confirmed PiD cases were available from the PIC, recruited from 31 international clinical or pathological research centres in the UK, France, Italy, Netherlands, Germany, Italy, Spain, Sweden, Australia, United States and Canada (**Supplementary Table 1)** Of the 321 cases, 151 were collected by the University College London (UCL) cohort and 170 by the Mayo Clinic Jacksonville (MCJ). For inclusion, all cases had to meet the strict PIC diagnostic criteria for PiD which have been described previously(11); as a minimum there needed to be the presence of Pick bodies with 3R tau positive and 4R tau negative inclusions. The additional presence of ballooned neurons and positive Gallyas staining was preferred to confirm diagnosis. All samples were screened for the known *MAPT* mutations so that these could be excluded in downstream analysis. The rationale for exclusion was that we wanted to look at genetic risk factors for apparently sporadic 3R tau pathology. Clinical and demographic data was collected for all cases, and included age at symptom onset, age at death and gender. This information was used to calculate the total disease duration, defined as age at death – age at symptom onset. Age at symptom onset was defined as the age at which first symptoms appeared, including initial cognitive dysfunction in judgment, language, or memory, or changes in behaviour or personality. Healthy controls with no clinical signs of neurological disease, a subset of whom had no pathology at post-mortem, were collected with the aim of having a ∼1:3 ratio (cases:controls), a similar age (defined by age at blood draw), and similar sex balance. Clinically defined controls were obtained from two sources; the Global Parkinson’s Genetics Program (GP2)(17) genotyped on the NBA, and the Invasive Fungal Infection and GENetics (IFIGEN) cohort(18) genotyped on the GSA array. A subset of 46 pathologically confirmed controls genotyped on the NBA were obtained from the Brains for Dementia Research (BDR) cohort.(19). The appropriate institutional review boards for each site approved the study, and written informed consent was obtained for each participant.

### Genotyping, Quality Control and Imputation

DNA was extracted using standard methods at the respective collection site (MCJ or UCL) as detailed in **Supplementary Methods A**. All samples from MCJ (North American samples), Sydney (Australian samples) and IFIGEN controls were genotyped by the local teams on the Illumina (Illumina, San Diego, CA, USA) Global Screening Array version 3 (GSA) (https://www.illumina.com/products/by-type/microarray-kits/infinium-global-screening.html). All UCL samples (European samples) and BDR control samples were genotyped on the Illumina NeuroBooster Array (NBA). Genotypes were called separately for each of the genotyping arrays using GenomeStudio version 2.0 (Illumina), based on the protocol published by Guo et al (20). All UCL cases and GP2 controls were screened for known *MAPT* mutations covered by the NBA and were excluded from downstream analysis if a known pathogenic *MAPT* mutation was identified. MCJ samples had already been screened for *MAPT* mutations before being included in the study.

Standard quality control procedures were performed in PLINK (v1.9)(21) and R (4.0.5, 2021-03-31) at the individual sample level and then the single nucleotide polymorphism (SNP) level. All the quality control (QC) steps detailed below were carried out separately for the NBA and GSA samples. Post-QC each dataset was imputed separately, then merged post-imputation based on overlapping SNPs for downstream analysis (further details in **Supplementary Methods A**).

### Candidate variant analysis

Specific variants that have previously been identified in related diseases were extracted pre-GWAS to check whether they showed any association with risk of PiD. This included variants identified in the primary tauopathies (PSP(8,22,23), CBD(24) and primary aged-related tauopathy (PART)(25) and clinically diagnosed FTD(26). We also checked for an association between *MAPT* haplotypes and risk of PiD, by extracting the six *MAPT* variants that define the H1-subhaplotype structure. In particular, we wanted to confirm the association of the H2 haplotype with risk of PiD that has been shown previously by directly genotyping the six *MAPT* sub-haplotype defining SNPs(11).

### Association analysis

The association between each variant and risk of PiD was examined using PLINK v1.9 to perform a logistic regression model that was adjusted for age, sex, genotype array, and three principle components (PC). Each variant was assessed under an additive model (i.e., number of minor alleles). Odds ratios (ORs) and 95% confidence intervals (CI) were estimated and correspond to each additional minor allele of the given variant. Due to the small sample size in comparison with standard GWAS cohorts, and limited clinical association studies on covariates of PiD, the model was chosen based on a stepwise logistic regression (*step* function in R *stats* package [version 3.6.2]). The full model included covariates: gender, genotype array, age and PC1-10 and the model selected based on minimising Akaike Information Criteria (AIC) and maximising the R^2^. Using this method, the final covariates used in the logistic GWAS were gender, genotype array, age (at death for cases and age at blood draw for controls), and three principal components (PC1, 8 and 10), which achieved the minimum AIC and maximum R^2^. A genome-wide significant threshold was defined at p < 5 × 10^-8^, with a threshold of p < 5 × 10^-6^ for a suggestive (nominal) association. Variant positions are reported on human genome version 37 (GRCh37/hg19).

### Genomic risk loci definition and gene mapping

Functional Mapping and Annotation of Genome-Wide Association Studies (FUMA) was used to annotate and functionally map the variants identified in the GWAS(27), defining genomic risk loci around variants with p<5 × 10^-6^, and including all variants correlated (R^2^ > 0.6) with the most significant variant. A conditional analysis was performed for the lead 5 loci identified with FUMA, conditioning on the lead SNP at each loci using CGTA-COJO (v.1.93.0 beta; https://yanglab.westlake.edu.cn/software/gcta)^86^, to confirm that there were no additional independent signals at each loci (**Supplementary Methods A**).

### Batch Effect Characterization and Sensitivity Analyses

We investigated potential confounding due to the genotyping array and age covariates, and the use of a stepwise method for covariate selection. To ensure the robustness of our findings, we conducted a series of additional diagnostic and sensitivity analyses. First, we conducted a control-control GWAS treating genotyping array as the phenotype to quantify systematic differences between NBA and GSA platforms. Second, we implemented three complementary analytical strategies: (1) a combined logistic regression including PC1, 2, and 3, 2) a combined logistic regression with ARRAY × AGE interaction term, (3) array-specific GWAS followed by fixed-effects meta-analysis. These are detailed in **Supplementary Methods B**.

### Fine-mapping and functional annotation

To nominate causal variants, fine-mapping was applied using SuSiE (v.0.12.27; https://github.com/stephenslab/susieR)(28) and FINEMAP (v.1.3.1;http://www.christianbenner.com/)(29) on all variants within 1Mb of the lead variant of each genomic risk loci. The *echolocatoR* R package (V. 1.4; https://github.com/RajLabMSSM/echolocatoR) was used to report the Union Credible Set SNPs (UCS), which is the union of all tool-specific CS_95%_, as well as the Consensus SNPs, which are those nominated from the two fine-mapping tools (further details in **Supplementary Methods A**). To further investigate cis and trans regulatory mechanisms in these nominated genomic regions, each locus was mapped to brain cell type specific enhancer-promoter interactome data, to regulatory elements data from the FANTOM5 (RRID:SCR_002678) project(30,31) M, and to functional DNA elements from the ENCODE dataset (RRID:SCR_006793, https://www.encodeproject.org/)(32) using the *echolocatoR* R package as detailed above.

### Colocalisation analysis

To investigate whether there was an overlap between the GWAS loci that reached nominal significance and expression quantitative trait loci (eQTLs), a colocalization analysis was performed using the *coloc* R package for all genes within ± 1Mb of the lead genomic loci SNP (version 5.1.0; https://cran.r-project.org/web/packages/colocr/index.html)(33). Given *coloc* calculates Bayes factors under the assumption that there is a single casual variant at a locus, we first performed conditional analysis, as detailed above, to confirm that there were no additional independent signals and so ensure that this assumption of a single casual variant was met. Further details are provided in **Supplementary Methods A**. In addition, to investigate whether the nominated loci regulate alternative splicing, a similar approach was followed using cortex splicing QTLs (sQTLs) from the GTEXv8(34) containing all variant-gene association from 255 individuals, based on LeafCutter (version 0.2.9 RRID:SCR_017639; https://davidaknowles.github.io/leafcutter/)(35)

### Assessment of gene transcript and protein expression of lead genes

Brain expression profiles of gene transcripts and encoded proteins highlighted by the GWAS were assessed, using a range of different publicly available online data sources (**Supplementary Methods A**).

## Results

### Cohort characterisation

321 autopsy confirmed PiD cases were considered for inclusion (**Supp. Table 1**). Of these, 143 cases were genotyped on the NBA (all from the University College London [UCL] cohort), and 178 cases were genotyped on the GSA (171 from the Mayo Clinic Jacksonville [MCJ] cohort and 7 from Sydney collected as part of the UCL cohort). 1446 controls were considered for inclusion; 989 from Global Parkinson’s Genetics Program (GP2)(36) that were genotyped on the NBA, and 457 from Invasive Fungal Infection and GENetics (IFIGEN) cohort(18) genotyped on the GSA. Samples excluded at each stage of the QC process are summarised in **Supp. Figure 1**. After quality control and filtering, 294 cases (135 NBA and 159 GSA), and 1055 controls (980 NBA and 75 GSA), covering 6,316,457 variants were available for association analysis. Due to the young average age of the GSA genotyped IFIGEN controls compared to the GSA genotyped PiD cases (Mean 37.7 vs 70.1 years), only GSA controls who were older than 50 years were selected for inclusion. This resulted in 75 GSA controls being selected (Mean 55.7 years) that were more closely matched in age, while still leaving enough GSA genotyped controls to allow inclusion of array type as a covariate to regress out array batch effects in the association analysis.

Demographics and basic clinical characteristics of the samples included for analysis after quality control are summarised in (**Table 1).** Overall, age (age at death for PiD cases and age at blood draw for controls) was slightly older for PiD cases compared to controls (Mean=69.4 years vs 66.4 years), and male sex was more common in PiD cases (63.2% vs. 36.7%). The mean age of onset for PiD cases was 58.9 years (SD = 8.0 years), mean age at death was 69.4 years (SD = 7.6 years), and correspondingly mean survival was 10.6 years (SD = 4.1 years). **Supp. Table 2** gives a breakdown of the clinical diagnoses for the 294 PiD cases; a total of 227 PiD cases (80.2%) had a clinical diagnosis of FTD (137 [46.5%] bvFTD, 60 [20.4%] PPA, and 39 [13.3%] not classified), 34 (11.6%) had a clinical diagnosis of AD, 14 (4.8%) had a clinical diagnosis of CBS, with the remainder being clinically diagnosed with vascular dementia, dementia not otherwise specified, or receiving no clinical diagnosis at all.

**Table 1.**
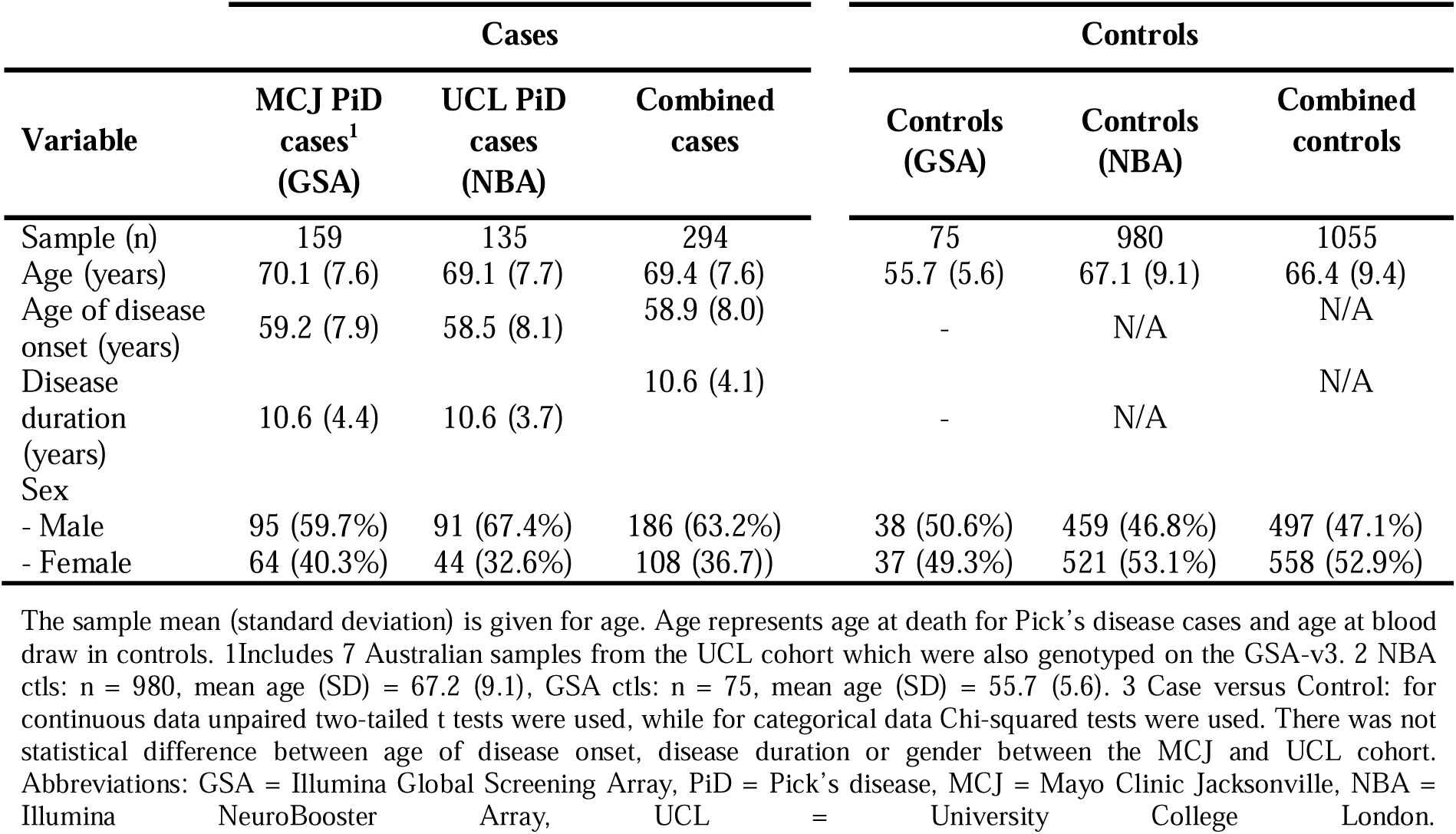
Clinical characteristics of samples included in GWAS.

| Variable | Cases |  |  | Controls |  |  |
| --- | --- | --- | --- | --- | --- | --- |
|  | MCJ PiD cases<sup>1</sup> (GSA) | UCL PiD cases (NBA) | Combined cases | Controls (GSA) | Controls (NBA) | Combined controls |
| Sample (n) | 159 | 135 | 294 | 75 | 980 | 1055 |
| Age (years) | 70.1 (7.6) | 69.1 (7.7) | 69.4 (7.6) | 55.7 (5.6) | 67.1 (9.1) | 66.4 (9.4) |
| Age of disease onset (years) | 59.2 (7.9) | 58.5 (8.1) | 58.9 (8.0) | - | N/A | N/A |
| Disease duration (years) | 10.6 (4.4) | 10.6 (3.7) | 10.6 (4.1) | - | N/A | N/A |
| Sex |  |  |  |  |  |  |
| - Male | 95 (59.7%) | 91 (67.4%) | 186 (63.2%) | 38 (50.6%) | 459 (46.8%) | 497 (47.1%) |
| - Female | 64 (40.3%) | 44 (32.6%) | 108 (36.7%) | 37 (49.3%) | 521 (53.1%) | 558 (52.9%) |

The sample mean (standard deviation) is given for age. Age represents age at death for Pick's disease cases and age at blood draw in controls. 1Includes 7 Australian samples from the UCL cohort which were also genotyped on the GSA-v3. 2 NBA ctls: n = 980, mean age (SD) = 67.2 (9.1), GSA ctls: n = 75, mean age (SD) = 55.7 (5.6). 3 Case versus Control: for continuous data unpaired two-tailed t tests were used, while for categorical data Chi-squared tests were used. There was not statistical difference between age of disease onset, disease duration or gender between the MCJ and UCL cohort. Abbreviations: GSA = Illumina Global Screening Array, PiD = Pick's disease, MCJ = Mayo Clinic Jacksonville, NBA = Illumina NeuroBooster Array, UCL = University College London.

All samples included from MCJ and Sydney had negative *MAPT* mutation screening. Details of 4 UCL cases, with predominant 3R tau pathology and concomitant *MAPT* mutations, are given in the **Supp. Results A**. 3 of these cases were excluded from the main analysis, while the Q230R was included given this is likely a benign polymorphism in *MAPT*.

### Targeted assessment of candidate variants

Before examination of the unbiased GWAS, we first assessed associations with risk of PiD for candidate variants that have previously been associated with tauopathies or related diseases (**Table 2**). After applying a Bonferroni correction for multiple testing (18 variants, P<0.0028 considered significant), our results confirmed the association between the *MAPT* H2 haplotype and PiD risk (OR: 1.52, 95% CI: 1.18-1.97, p=0.001) that has been previously demonstrated through direct genotyping of rs8070723 in previous work from our group(11). Analysis of MAPT H1 and H2 haplotype frequency showed an increase (Chi square: _χ_ = 6.04, df =2, p = 0.003) in both H1/H2 heterozygotes (45.6% PiD cases vs. 36.1% controls) and H2/H2 homozygotes (6.8% PiD cases vs. 5% controls) (**Supp. Table 3**). There was 100% concordance between the direct genotyping and chip-based imputation of rs8070723 (H2 tagging variant) (data not shown). Of the other 17 variants tested, none passed the analysis-wide significance threshold, though *MOBP* was associated with risk of PiD at the p <0.05 level (OR: 0.76, 95% CI: 0.59 – 0.98, p = 0.03).

**Table 2.** Candidate variant analysis using GWAS data.

| Disease | Chr | SNP | Nearest gene | MAF<br>(cases) | MAF<br>(controls) | MAF<br>(total cohort) | OR (95% CI) | p value |
| --- | --- | --- | --- | --- | --- | --- | --- | --- |
| *MAPT* (H2) | 17 | rs8070723 | *MAPT* | 0.29 | 0.23 | 0.24 | 1.52 (1.18 - 1.97) | 0.001<sup>a</sup> |
| *MAPT* (H1c) | 17 | rs242557 | *MAPT* | 0.35 | 0.34 | 0.34 | 1.02 (0.80 - 1.20) | 0.86 |
| AD | 19 | rs429358 | *ApoE* | 0.15 | 0.13 | 0.13 | 1.40 (1.02 - 1.93) | 0.04 |
| AD | 19 | rs7412 | *ApoE* | 0.09 | 0.09 | 0.09 | 0.97 (0.66 - 1.43) | 0.89 |
| PSP | 1 | rs564309 | *TRIM11* | 0.09 | 0.09 | 0.09 | 1.31 (0.88 - 1.96) | 0.19 |
| PSP | 12 | rs2242367 | *SLC2A13* | 0.29 | 0.26 | 0.27 | 1.11 (0.86 - 1.42) | 0.43 |
| PSP | 1 | rs1411478 | *STX6* | 0.42 | 0.41 | 0.41 | 1.08 (0.86 - 1.35) | 0.51 |
| PSP | 3 | rs1768208 | *MOBP* | 0.25 | 0.28 | 0.28 | 0.76 (0.59 - 0.98) | 0.03<sup>b</sup> |
| PSP | 2 | rs7571971 | *EIF2AK3* | 0.28 | 0.26 | 0.26 | 0.96 (0.74 - 1.25) | 0.77 |
| FTD-TDP | 7 | rs1990622 | *TMEM106B* | 0.42 | 0.39 | 0.40 | 1.24 (0.98 - 1.56) | 0.07 |
| FTD | 11 | rs302668 | *RAB38* | 0.37 | 0.35 | 0.35 | 1.15 (0.90 - 1.45) | 0.26 |
| FTD | 11 | rs16913634 | *RAB38/CTSC* | 0.09 | 0.09 | 0.09 | 0.95 (0.63 - 1.45) | 0.81 |
| FTD | 6 | rs9268877 | *HLA-DRA/DRB5* | 0.42 | 0.44 | 0.44 | 1.04 (0.83 - 1.31) | 0.72 |
| FTD | 6 | rs9268856 | *HLA-DRA/DRB5* | 0.25 | 0.24 | 0.24 | 0.91 (0.70 - 1.19) | 0.50 |
| FTD | 6 | rs1980493 | *BTNL2* | 0.15 | 0.14 | 0.14 | 1.04 (0.74 - 1.46) | 0.82 |
| PART | 4 | rs56405341 | *JADE1* | 0.27 | 0.29 | 0.28 | 1.01 (0.79 - 1.31) | 0.91 |
| CBD | 2 | rs963731 | *SOS1* | 0.05 | 0.05 | 0.05 | 0.63 (0.37 - 1.08) | 0.09 |
| CBD | 8 | rs643472 | *lnc-KIF13B-1* | 0.23 | 0.22 | 0.27 | 1.13 (0.74 - 1.25) | 0.35 |

Logistic regression additive model adjusted for gender, age, array and 3 PCs (PC1, 8 and 10) were used to study the association of candidate loci with risk of PiD in the total cohort (294 cases, 1055 controls). a significant after correction for multiple comparisons (Bonferroni: $p=0.05/18=0.0028$ ). b not significant after correction for multiple comparisons. Abbreviations: Chr = chromosome, CI = confidence interval, MAF = minor allele frequency, OR = odds ratio.

### Association Results

Using a case-control logistic regression model, adjusting for gender, genotype array, age and three genetic principal components (PCs 1, 8,10) to account for population substructure, we assessed the role of 6,316,457 variants on the risk of developing PiD. The genomic inflation factor (λ) was 0.99 (λ_1000 = 0.97) (**Supp Figure 2a**) demonstrating no confounding by population stratification. No disease-associated variants reached genome-wide significance (*p* < 5 × 10^-8^), but there were suggestive associations (defined as *p* < 5 × 10^-6^) at five genomic loci (**Figure 1**), with the lead SNP at each locus shown in **Table 3**. The lead locus was on Chromosome 4 (rs11216197, OR = 7.53, 95% Confidence Interval (CI) = 3.62-15.65, p = 6.37 × 10^-8^), with the second locus on chromosome 11 (rs66481907, OR = 2.10, 95% CI = 1.54-2.84, *p* = 1.83 × 10^-6^). **Figure 2** shows more detailed regional association plots for each of the five genomic loci with suggestive associations. Conditional analyses performed on the lead variant at each of these five loci confirmed that there were no additional independent signals (**Supp. Figure 3**).

**Figure 1.**
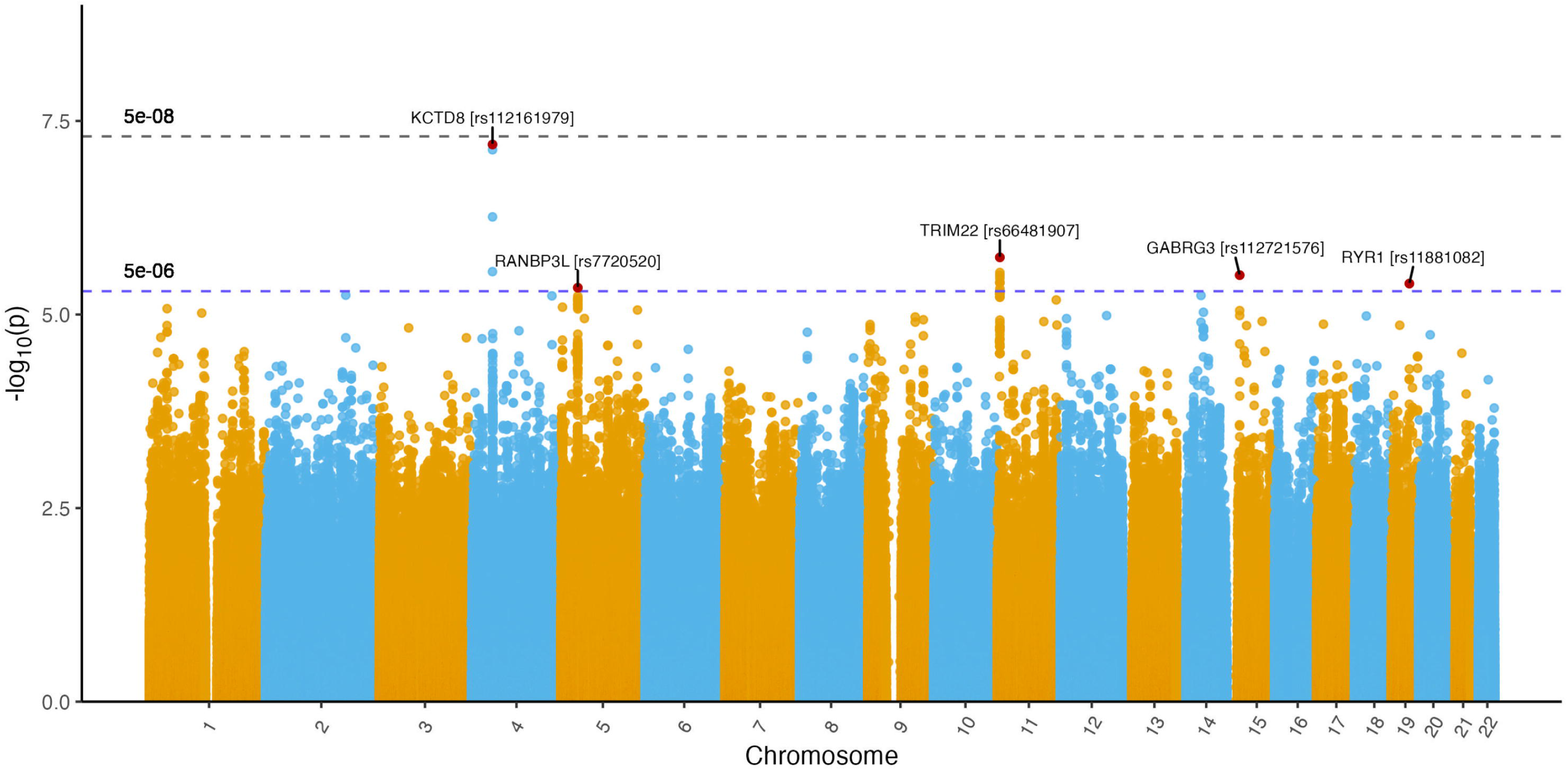
Association plot for PiD. Manhattan plot showing -log10 (P) values from logistic regression of imputed variants corrected for age, gender, array and three principal components (PC1, 8 and 10). Red dots indicate the variant (rsID and nearest gene labelled) with the lowest p value at each genomic locus that reached nominal significance (p < 5 × 10-6) indicated by the blue dashed line. Genome-wide significance was set at p < 5 × 10-8 and indicated by the grey dashed line

**Figure 2.**
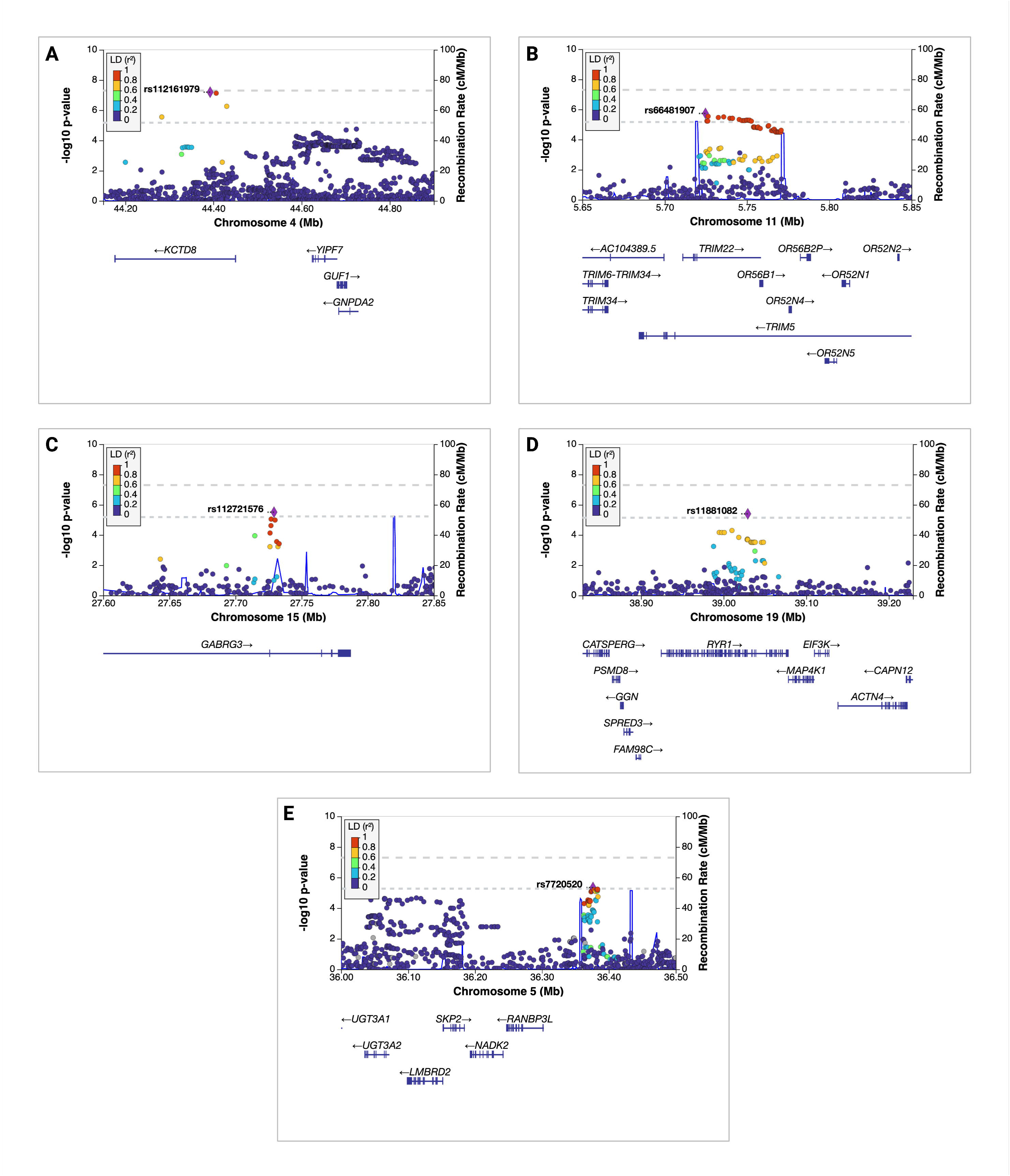
Regional association plots and recombination rates at suggestive genomic loci. (A-E) (A) Regional association plots at 4: 44392571 (rs112161979)), (B) 11: 5724803 (rs66481907), (C) 15: 27729149 (rs112721576), (D) 19:39029201 (rs11881082), and (E) 5:36376351 (rs7720520). The index variants are indicated by a purple diamond and corresponding rsID. Linkage disequilibrium between the index variant and nearby variants, as measured by r2, is colour-coded (dark blue: 0 ≤ r2 < 0.2; light blue: 0.2 ≤ r2 < 0.4; green: 0.4 ≤ r2 < 0.6; orange: 0.6 ≤ r2 < 0.8; red: 0.8 ≤ r2 ≤ 1; blue: no r2 available). Genome-wide significance was set at *p* < 5 × 10-8 and indicated by the top grey dashed line, while nominal significance (p < 5 × 10-6) is indicated by the lower grey dashed line All plots were generated in http://locuszoom.org/.

**Table 3.** Top independent SNPs at suggestive loci from PiD GWAS.

| Chr | BP | SNP | Nearest gene | Minor allele | MAF |  |  |  |  |
| --- | --- | --- | --- | --- | --- | --- | --- | --- | --- |
|  |  |  |  |  | Cases | Controls | NFE* | OR (95% CI) | p value |
| 4 | 44,392,571 | rs112161979 | *KCTD8* | A | 0.040 | 0.013 | 0.016 | 7.53 (3.62-15.65) | 6.37 x 10<sup>-8</sup> |
| 11 | 5,724,803 | rs66481907 | *TRIM22* | A | 0.206 | 0.126 | 0.120 | 2.10 (1.54-2.84) | 1.83 x 10<sup>-6</sup> |
| 15 | 27,729,149 | rs112721576 | *GABRG3* | G | 0.045 | 0.029 | 0.038 | 3.73 (2.14-6.48) | 3.10 x 10<sup>-6</sup> |
| 19 | 39,029,201 | rs11881082 | *RYR1* | A | 0.070 | 0.050 | 0.053 | 2.96 (1.87-4.69) | 4.00 x 10<sup>-6</sup> |
| 5 | 36,376,351 | rs7720520 | *RANBP3L* | G | 0.421 | 0.328 | 0.337 | 1.76 (1.38-2.23) | 4.50 x 10<sup>-6</sup> |

ORs, 95% CIs, and p-values result from logistic regression models that were adjusted for age, sex, and PCs 1, 8, and 10. ORs correspond to each additional minor allele of the given variant
\*Non-Finnish Europeans (GnomAD v2.1.1). Abbreviations: BP = base-pair coordinate, Chr = chromosome, CI = confidence interval, MAF = minor allele frequency, OR = odds ratio.

To ensure that our GWAS results were robust and not confounded by technical or cohortLJspecific effects, we performed comprehensive batch effect characterization and sensitivity analyses as detailed in **Supp Materials B**. Performing the GWAS with PC1-3 instead of the stepwise regression selection of covariates (PC1,8,10) confirmed the signal at our lead loci; rs11216197 (OR 6.83, 95% CI 3.32-14.06, p = 1.82 × 10^-7^) and rs66481907 (OR 2.13, 95% CI 1.57-2.89, p = 1.09 × 10^-6^). The GWAS with addition of an *Age:Array* interaction term also confirmed the signal at these loci with similar effect sizes albeit with a slightly reduced *p* value (rs11216197; OR 5.31, 95% CI 2.55LJ11.06, p = 4.57×10LJLJ, rs66481907; OR 2.13, 95% CI 1.57-2.89, p = 5.36 × 10^-6^ ). Meta-analysis of the summary statistics for the GWAS performed on the GSA and NBA array separately confirmed the signal at our lead loci rs11216197 (OR 5.31, 95% CI 2.55 - 11.06, *p* = 4.57 × 10^-6^) and rs66481907 (OR 1.90, 95% CI 1.37-2.64, *p* = 1.38 × 10^-4^), but with an attenuated level of significance due to the reduced statistical power inherent to meta-analysis (**Supp. Figure 4)**. As these complimentary approaches did not fundamentally alter the finding of a large effect size (**Supp. Table 4**) and the combined analysis is better powered to detect associations, as shown by the comparative QQ plots (**Supp. Figure 2)**, we proceeded with the results from our primary combined model.

The most significant SNP, rs11216197 on Chromosome 4, is an intronic variant located in *KCTD8* (**Figure 2A**). The *KCTD8* gene encodes a potassium tetramerisation domain that facilitates GABA_B_ receptor expression in axonal terminals and contributes to presynaptic excitation by GABA_B_ receptors(37,38). Approximately 500kb downstream of the lead variant there are three genes: *YIPF7, GUF1* and *GNPDA2*. rs112161979 is an expression quantitative trait locus (eQTL) for *GNPDA2* in blood (GTEXv8; *p* = 6.4 × 10^-5^) and tibial nerve tissue (GTEXv8; *p* = 6.3 × 10^-5^), and for *GUF1* in blood (GTEXv8; *p* = 1.9 × 10^-4^), though not for either gene in the brain.

The next suggestive association was on Chromosome 11; the lead variant in this region was rs66481907 (OR = 2.10, 95% CI = 1.54-2.84, *p* = 1.83 × 10^-6^), an intronic SNP located in the *TRIM22* gene (**Figure 2B**). TRIM22 is a member of the tripartite motif-containing (TRIM) superfamily, all of which have an E3 ubiquitin ligase function, and are involved in a wide range of cellular processes including degradation of misfolded proteins(39), regulation of the NLRP3 inflammasome signalling pathway(40), and antiretroviral activity against a wide range of viruses including HIV, Influenza A, Hepatitis B and C(41) playing an important role in the innate immune response to infection(42). Interestingly the rs66481907 SNP is a sQTL for *TRIM22* in nerve-tibial tissue (GTEXv8; intron id 5708603:5709053:clu_7256, *p* = 1.2 × 10^-8^), is associated with both non-coding transcripts with a retained intron, as well transcripts targeted for nonsense mediated decay in ENSEMBL (RRID:SCR_002344; ENSEMBL) and has a Combined Annotation Dependent Depletion (CADD) score of 10.02 placing it in the top 10% most deleterious variants in the genome.

The final three suggestive genomic loci with variants showing nominal significance were Chromosome 15 (lead SNP rs112721576, an intronic variant in *GABRG3* (OR = 3.73, 95% CI = 2.14-6.48, 3.10 × 10^-6^)), Chromosome 19 (lead SNP rs11881082, a splice site variant in *RYR1* (OR = 2.96, 95% CI = 1.87-4.69, 4.00 × 10^-6^), and Chromosome 5 (lead SNP rs7720520, an intergenic variant close to *RANBP3L* (OR = 1.

### Fine-mapping, colocalisation and transcript expression

Under the single causal variant assumption (supported by the conditional analysis) statistical fine-mapping was performed at the two leading genomic loci (Chromosome 4 and 11) with FINEMAP(29) and SuSiE(28). No consensus causal SNPs (supported by both fine-mapping techniques) were identified at the lead locus (Chromosome 4). However, SuSiE nominated rs990356 as the likely causal SNP (posterior probability 1), a 3’ UTR variant in *YIPF7* (Yip1 Domain Family Member 7) (**Supp. Table 5 and Supp. Figure 5A**). Mapping the Chromosome 4 locus against genomic regulatory elements did not show any significant signals. Fine-mapping of the Chromosome 11 locus also failed to demonstrate a consensus causal SNP across the two fine-mapping algorithms. SuSiE nominated three SNPs as potentially causal, while FINEMAP nominated one (**Supp. Table 5 and Figure 5B**). Of particular interest was the rs7397032 SNP identified by SuSiE; this is in high linkage disequilibrium (LD) (D’ 0.95, R^2^ 0.86) with the lead SNP from the GWAS (rs66481907), is a 3’UTR variant in *TRIM22*. Mapping this region against available genomic regulatory elements demonstrated that the lead SNP, and surrounding SNPs in high LD, sit within a conserved transcription factor binding site, supporting that this locus is involved in transcriptional regulation of surrounding genes (Bottom panel in **Figure 5B**)

To further delineate the effects of the chromosome 4 and 11 loci on regulation of gene expression, colocalisation analysis was performed using cortical cis-eQTLs from the MetaBrain dataset(29). There was no evidence of colocalisation (defined as a PP.H4 of > 0.85) between the two lead PiD GWAS loci and eQTLs for genes within ± 1Mb of the locus. At the chromosome 4 locus *KCTD8* had a PP.H4 of colocalisation of 0.02, while the three genes downstream *YIPF7, GUF1* and *GNPDA2* had PP.H4s of 0.74, 0.81 and 0.72 respectively. At the chromosome 11 locus, *TRIM22* and *TRIM5* both had PP.H4s of 0.04. Given the suggestion that the causal SNP for the chromosome 4 signal may be mediated by rs990356 in the 3’ region of *YIPF7*, regional association plots for the eQTL signal from *KCTD8* and *YIPF7* and the PiD GWAS signal were plotted (**Figure 3**). Visual inspection of these plots demonstrates that the GWAS signal is more closely aligned with the *YIPF7* than the *KCTD8* eQTL signal suggesting that the GWAS signal at the chromosome 4 locus could be mediated by dysregulation of *YIPF7* gene expression. However, this association did not meet the predefined threshold of certainty for colocalisation and so with the current sample size this cannot be confirmed. Given the chromosome 11 lead SNP (rs66481907) is a sQTL for *TRIM22* in nerve-tibial tissue, we explored whether this region could have a role in alternative splicing of the gene in cortical tissue, so performed colocalisation analysis using cortex sQTLs from the GTExv8(28). Again, there was no evidence for colocalisation of the GWAS signal in this region and sQTLs for *TRIM22* in cortical tissue (PP.H4: min 0.036, max 0.055).

**Figure 3.**
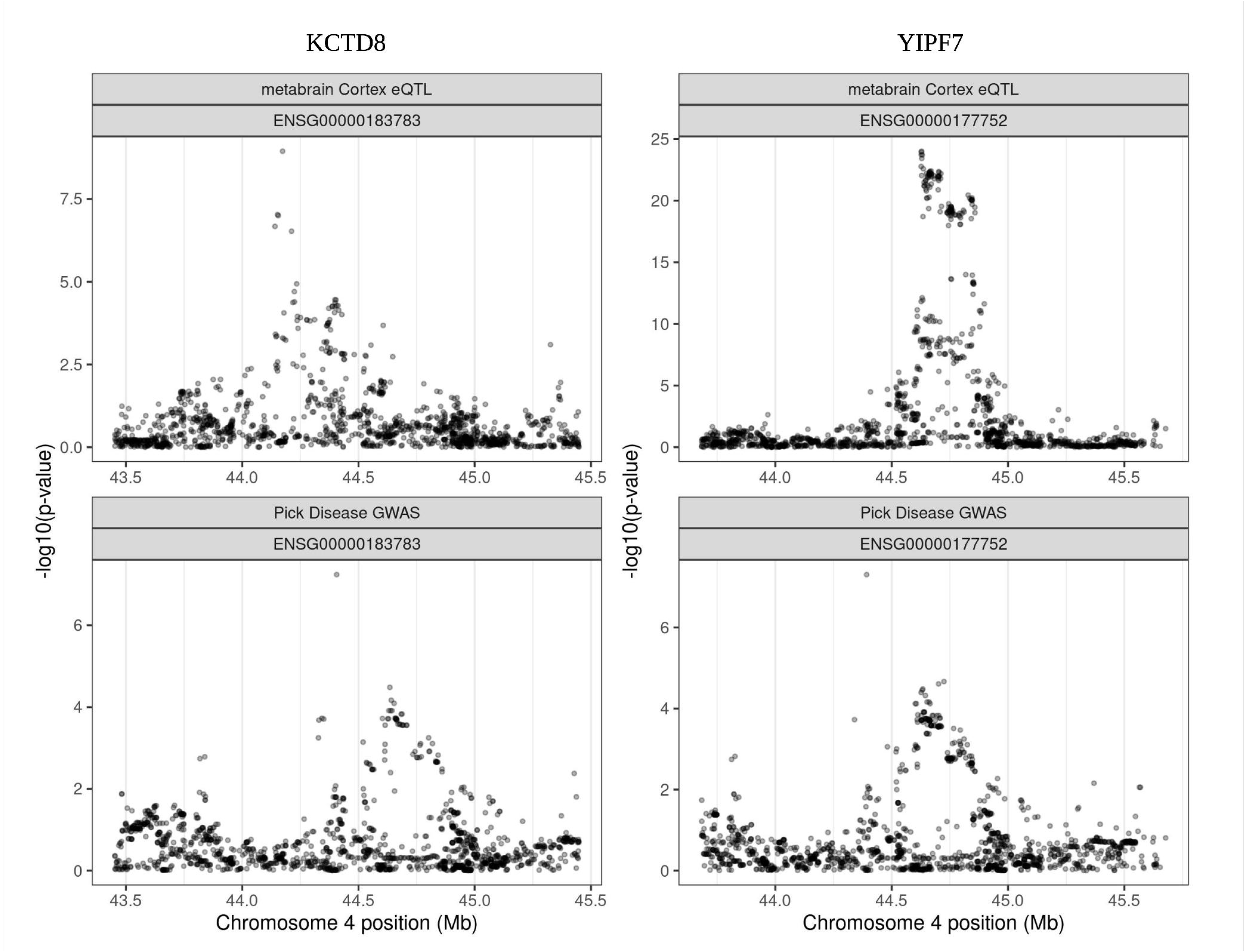
Regional plots from PiD GWAS and MetaBrain cis-eQTLs for KCTD8 and YIPF7 genes. The PP.H4 of there being a shared causal variant associated with both PiD (bottom panels) and regulation of gene expression (top panels) was 0.02 for KCTD8 (left side) and 0.74 for YIPF7 (right side).

Given the suggestion from the fine-mapping and colocalisation analysis that the causal SNP at the chromosome 4 locus was located in YIPF7, we investigated the gene and protein expression profile of this gene in addition to KCTD8, alongside TRIM22 located at the chromosome 11 locus (**Supp. Figures 6-8**). Full details of these analyses are provided in **Supplementary Methods and Results A**.

## Discussion

Using 294 autopsy-confirmed PiD cases collected through the PIC, we conducted a GWAS to identify genetic risk factors for the disease. No variant reached genome-wide significance, after exclusion of three cases with MAPT mutations. A previous GWAS of 219 autopsy-confirmed corticobasal degeneration (a 4R tauopathy) did identify significant common variants (24), which suggests that PiD risk either lacks significant genetic risks beyond MAPT, or has genetic modifiers of smaller effect size than this study was powered to detect. Five loci showed suggestive associations, with the strongest signals in *KCTD8* on chromosome 4 (rs112161979, p = 6.37 × 10LJLJ) and *TRIM22* on chromosome 11 (rs66481907, p = 1.83 × 10LJLJ). Sensitivity analyses indicated that 4 of the 5 associations at *KCTD8*, *TRIM22*, *RANBP3L*, and *GABRG3* are robust and reproducible with stable effect sizes.

The association of the MAPT H2 haplotype with PiD, previously established through direct genotyping, was confirmed (11). Here, with genome-wide data, population stratification could be accounted for, and the association strengthened (OR 1.52, 95% CI 1.18–1.97 vs. OR: 1.35, 95% CI: 1.12-1.64, *p* = 0.0021 respectively). Variants associated with PSP and CBD were not associated with PiD, except for a nominal signal at MOBP (OR 0.76, 95% CI 0.59 - 0.98, p = 0.03), that did not survive multiple testing and showed the opposite direction of effect, supporting distinct genetic architectures and pathophysiological mechanisms between 3R and 4R tauopathies.

The lead signal at the chromosome 4 locus comes from an intronic SNP located within the *KCTD8* gene. There are no deleterious coding variants in LD with the lead SNP, no colocalisation of the GWAS signal in this region with brain eQTLs of *KCTD8* or the three genes c.500kb downstream (*YIPF7, GNPDA2* or *GUF1*), and fine-mapping was inconclusive with regards to a consensus causal SNP. However, given the colocalisation analysis was likely to be underpowered due to the absence of genome-wide significance in the GWAS data, we cannot exclude that this variant does not actually affect expression of these downstream genes. *YIPF7* is an interesting potential candidate gene at this locus given its predicted interactions with proteins that when mutated are known to cause both neurodevelopmental disorders as well as ALS. The predicted interaction with the DM1 gene *DMPK* is also intriguing given the known presence of 3R tau at post-mortem in these patients(43).

The *KCTD8* gene is highly expressed within the brain, predominantly in neurons and oligodendrocytes, and the protein it encodes has been shown, using immunohistochemistry, to be present in the cortex. scRNA analysis suggests its expression is specifically enriched in habenula neurons, which is supported by in situ hybridisation analysis of *KCTD* receptor transcripts in the mouse brain showing its particular abundance in the medial habenula followed by the subiculum of the hippocampus(44). The habenula and the subiculum are of interest with regards to PiD. The habenula is affected by neurodegeneration in behavioural variant FTD (bvFTD) (the most common presentation of PiD) showing a 29% lower volume compared to controls(45), and its degeneration can lead to perseveration or disinhibition and impulsivity(46), symptoms commonly seen in bvFTD. The subiculum is commonly affected by Pick’s pathology with high densities of Pick bodies found in this part of the hippocampus at post-mortem(47). The *KCTD* family of proteins are currently poorly characterised, though are increasingly recognised to be involved in a range of neurocognitive, neurodevelopmental and neuropsychiatric disorders. These include mutations in *KCTD3* (global developmental delay, seizures and cerebellar hypoplasia)(48,49), *KCTD13* (autism and schizophrenia)(50–52), *KCTD17* (myoclonus-dystonia)(53–55), *KCTD12* (bipolar 1 disorder)(56), and *KCTD7* which can cause either a severe progressive myoclonic epilepsy syndrome (EPM3)(57) or neuronal ceroid lipofuscinosis(58) depending on the specific mutation. *KCTD8*, specifically, acts as an auxiliary subunit of GABA_B_ receptors and has been shown to facilitate their axonal expression in habenula cholinergic neurons(37). GABA deficits in FTD have long been recognised, with evidence that GABAergic neurons are markedly reduced in FTD at post-mortem(59), with PET(60) and MRS imaging studies demonstrating GABAergic deficits in vivo(61). Overall, the localisation of the lead SNP, and the gene expression profiles described above make *KCTD8* the most plausible candidate gene at the chromosome 4 locus, though further work will be needed both to validate this signal, and also delineate the mechanisms by which it contributes to PiD pathology.

The GWAS signal at chromosome 11 implicates *TRIM22*. The lead SNP rs66481907 is a sQTL in nerve tibial tissue for *TRIM22*, is located in alternatively spliced transcripts with retained introns (non-functional) and others targeted for nonsense mediated decay, and has a CADD score of 10.02 placing it in the top 10% most deleterious variants in the genome. *TRIM22* is expressed both at the transcript and protein level in the brain and is enriched within microglia in contrast to *KCTD8* which is predominantly expressed in neurons. The TRIM family of proteins, the majority of which have E3 ubiquitin ligase activity, have a wide range of functions within cellular processes including eliminating misfolded proteins (via autophagy)(62,63), the ubiquitin proteosome system (UPS)(39,64), and endoplasmic-reticulum associated degradation (ERAD)(65), antiviral activity(66) and regulation of the NF-kB/NLRP3 inflammasome pathway(67). Mutations in TRIMs are increasingly recognised as a cause of a wide range of diseases including a more aggressive phenotype in PSP (*TRIM11/17*)(23), cerebral small vessel disease (*TRIM47*) (68), and limb girdle muscular dystrophies (*TRIM 32*)(69). Although *TRIM22* was first identified through its anti-viral properties in HIV infection(70), more recent work has demonstrated its role in autophagy through interaction with autophagy regulators ULK1 and Beclin1(62,63), as well as effectively promoting elimination of misfolded proteins via the UPS during cell transformation(71). Consistent with this role is the finding that *TRIM19/PML*, which promotes clearance of misfolded proteins (including ataxin-7 in SCA7) via the proteosome(72), colocalises with *TRIM22* in nuclear bodies under IFN-y stimulation(73). Overall, *TRIM22* is a biologically plausible candidate gene for risk of PiD, based on the hypothesis that variation at this locus modifies the protein function (potentially through nonsense mediate decay or alternative splicing of gene transcripts), leading to decreased degradation of toxic 3R tau protein via the UPS and / or the autophagy pathway(s).

There are several limitations to this study. Ideally, a GWAS should include a discovery phase identifying genome-wide significant signals, followed by an independent replication phase. Despite assembling essentially all autopsy-confirmed PiD cases worldwide, the sample size remains small and a two-stage design was therefore not feasible. Estimated accrual through the PIC network is only 10– 15 new autopsy-confirmed cases per year, and without pathology-specific in-vivo biomarkers to distinguish PiD from 4R tauopathies and TDP-43 disease, imminent targeted enrichment of clinical cohorts remains unrealistic. Thus, replication of these findings may not be possible in the near term. Nonetheless, we hope the PIC facilitates coordinated future case collection under standardised pathological criteria and stimulates functional and biomarker-development work. A further limitation is that, aside from direct *MAPT* haplotype genotyping, lead SNPs at the suggestive loci were imputed. Although stringent quality control thresholds were applied, small inaccuracies in allele frequency estimation can disproportionately affect effect size and p-value, particularly for rare variants. This is salient at the chromosome 4 locus where the lead allele is present at low frequency. Future work should involve direct sequencing of these regions to confirm genotypes and determine whether the GWAS signal reflects, or tags, a deleterious rare variant.

In conclusion we have performed the first GWAS with the aim of identifying the genetic drivers of disease risk in PiD. The data confirms that the *MAPT* H2 haplotype is associated with PiD, as opposed to the more common H1 haplotype in PSP and CBD. Known risk variants for the 4R tauopathies are not associated with disease, which suggests that the underlying genetic architecture of disease risk for PiD is distinct. This has important implications for the future development of therapeutics to treat PiD, and emphasises the need for PiD specific biomarkers to identify these individuals in life. *KCTD8*, the most plausible gene at the lead locus, modulates GABA_B_ receptor expression within anatomically relevant regions of the brain and implicates dysregulation of the GABAergic neurons as an important driver of disease pathology. This is supported by the other suggestive association within the *GABRG3* gene (a GABA_A_ receptor subunit) on chromosome 15. In addition, common variation in *TRIM22* may also play a role in disease pathogenesis, potentially through perturbation of the UPS and its ability to eliminate toxic tau species, representing a potential target for disease modifying therapies. Future work should focus on further GWASs with larger sample sizes to confirm or refute the findings from this study, whole genome sequencing of the lead loci to identify if rare deleterious variants are driving the signals here, and functional studies, ideally in induced pluripotent stem cells, to reveal how these genes may be contributing to PiD risk and so better elucidate the pathogenic pathway resulting in 3R tau accumulation.

## Data Availability

All data produced in the present study are available upon reasonable request to the authors

## Acknowledgements

Funding for main authors listed below. For PIC members names and funding including brainbank acknowledgments please see **Supplementary Materials (E)**.

WJS receives a Wellcome Trust Clinical PhD Fellowship (220582/Z/20/Z and personal funding from the Rotha Abraham Trust. WJS would like to thank the Rotha Abraham Trust and The Mason Legacy bequest for ongoing support of his work in neurodegenerative disease. RR is funded by ASAP. TL receives an Alzheimer’s Research UK senior fellowship. JBR is supported by the Medical Research Council (MC_UU_00030/14; MR/T033371/1), the Wellcome Trust (220258), the NIHR Cambridge Biomedical Research Centre (NIHR203312), the Cambridge Centre for Parkinson-plus and Evelyn Trust. The views expressed are those of the authors and not necessarily those of the NIHR or the Department of Health and Social Care. HRM is supported by research grants from Parkinson’s UK, Cure Parkinson’s Trust, PSP Association, CBD Solutions, Drake Foundation, Medical Research Council, and Michael J Fox Foundation. JDR receives a Miriam Marks Brain Research UK Senior Fellowship and has received funding from an MRC Clinician Scientist Fellowship (MR/M008525/1) and the NIHR Rare Disease Translational Research Collaboration (BRC149/NS/MH). OAR and DWD are both supported by National Institute of Neurological Disorders and Stroke (NINDS) Tau Center without Walls Program (U54-NS100693) and NIH (UG3-NS104095). DWD receives research support from the NIH (P30 AG062677; U54-NS100693; P01-AG003949), CurePSP, the Tau Consortium, and the Robert E. Jacoby Professorship. OAR is supported by NIH (P50-NS072187; R01- NS078086; U54-NS100693; U54- NS110435), Department of Defence (DOD) (W81XWH-17- 1-0249) The Michael J. Fox Foundation, The Little Family Foundation, the Mangurian Foundation Lewy Body Dementia Program at Mayo Clinic, the Turner Family Foundation, Mayo Clinic Foundation, and the Center for Individualized Medicine. Mayo Clinic is also an LBD Center without Walls (U54-NS110435). MS and JH wish to thank Fidelty Bermuda Foundation and Dolby Family trust for their continued support of our work in genetics of neurodegenerative diseases.

## Author contributions

WJS, RRV, JDR, JH, OAR and MS were involved in conceptualisation and design of the study. WJS, RRV, NT, HLM, TL, SFR and DD were involved in IHC of PiD brains, extracting DNA and genotyping the samples. WJS, AMC, RR, KM and MS carried out the formal genetic and computational analysis. WJS, MS, VEP and MGH designed and implemented the statistical analysis. WJS, RRV and MS drafted the manuscript. All authors were involved in funding, acquisition, resources, validation, critically reviewing and approving the final version of the manuscript. All PIC members were involving in acquisition of samples, resources, reviewing and approving the final manuscript.

## References

1. Shi Y, Zhang W, Yang Y, Murzin AG, Falcon B, Kotecha A, et al. Structure-based classification of tauopathies. Nature. 2021;598(7880):359–63.

2. Falcon B, Zhang W, Murzin AG, Murshudov G, Garringer HJ, Vidal R, et al. Structures of filaments from Pick’s disease reveal a novel tau protein fold. Nature. 2018 Sep;561(7721):137–140

3. Goedert M. Tau filaments in neurodegenerative diseases. FEBS Lett. 2018;592(14):2383–91.

4. Ghetti B, Oblak AL, Boeve BF, Johnson KA, Dickerson BC, Goedert M. Invited review: Frontotemporal dementia caused by microtubule-associated protein tau gene (MAPT) mutations: a chameleon for neuropathology and neuroimaging Frontotemporal dementia caused by microtubule-associated protein tau gene (MAPT) mutations: chameleon for neuropathology and neuroimaging. Neuropathol Appl Neurobiol. 2015;41:24–46.

5. Schweighauser M, Garringer HJ, Klingstedt T, Nilsson KPR, Suzukake MM, Murrell JR, et al. Mutation Δ K281 in MAPT causes Pick’s disease. 2023 Aug;146(2):211–226.

6. Stefansson H, Helgason A, Thorleifsson G, Steinthorsdottir V, Masson G, Barnard J, et al. A common inversion under selection in Europeans. Nat Genet. 2005;37(2):129–37.

7. Baker M, Litvan I, Houlden H, Adamson J, Dickson D, Perez-Tur J, et al. Association of an extended haplotype in the tau gene with progressive supranuclear palsy. Hum Mol Genet. 1999;8(4):711–5.

8. Höglinger GU, Melhem NM, Dickson DW, Sleiman PMA, Wang LS, Klei L, et al. Identification of common variants influencing risk of the tauopathy progressive supranuclear palsy. Nat Genet. 2011 Jun 19;43(7):699–705.

9. Houlden H, Baker M, Morris HR, MacDonald N, Pickering-Brown S, Adamson J, et al. Corticobasal degeneration and progressive supranuclear palsy share a common tau haplotype. Neurology. 2001 Jun 26;56(12):1702–6.

10. Kouri N, Ross OA, Dombroski B, Younkin CS, Serie DJ, Soto-Ortolaza A, et al. Genome-wide association study of corticobasal degeneration identifies risk variants shared with progressive supranuclear palsy. Nat Commun. 2015 Jun 16;6.

11. Valentino RR, Scotton WJ, Roemer SF, Lashley T, Heckman MG, Shoai M, et al. MAPT H2 haplotype and risk of Pick’s disease in the Pick’s disease International Consortium: a genetic association study. Lancet Neurol. 2024;23(5):487–99.

12. Morris HR, Baker M, Yasojima K, Houlden H, Khan MN, Wood NW, et al. Analysis of tau haplotypes in Pick’s disease. Neurology. 2002;59(3):443–5.

13. Russ C, Lovestone S, Baker M, Pickering-Brown SM, Andersen PM, Furlong R, et al. The extended haplotype of the microtubule associated protein tau gene is not associated with Pick’s disease. Neurosci Lett. 2001;299(1–2):156–8.

14. Sanchez-Contreras MY, Kouri N, Cook CN, Serie DJ, Heckman MG, Finch NA, et al. Replication of progressive supranuclear palsy genome-wide association study identifies SLCO1A2 and DUSP10 as new susceptibility loci. Mol Neurodegener. 2018 Dec 9;13(1):37.

15. Jabbari E, Woodside J, Tan MMX, Shoai M, Pittman A, Ferrari R, et al. Variation at the TRIM11 locus modifies progressive supranuclear palsy phenotype. Ann Neurol. 2018;84(4):485–96.

16. Jabbari E, Koga S, Valentino RR, Reynolds RH, Ferrari R, Tan MMX, et al. Genetic determinants of survival in progressive supranuclear palsy: a genome-wide association study. Lancet Neurol. 2021 Dec;20(2):107–16.

17. Parkinson TG, Program G. GP2: The Global Parkinson’s Genetics Program. Movement Disorders. 2021;36(4):842–51.

18. Doni A, Parente R, Laface I, Magrini E, Cunha C, Colombo FS, et al. Serum amyloid P component is an essential element of resistance against Aspergillus fumigatus. Nat Commun. 2021 Jun 18;12(1):3739.

19. Francis PT, Hayes GM, Costello H, Whitfield DR. Brains for Dementia Research: The Importance of Cohorts in Brain Banking. Neurosci Bull. 2019 Apr;35(2):289–294.

20. Guo Y, He J, Zhao S, Wu H, Zhong X, Sheng Q, et al. Illumina human exome genotyping array clustering and quality control. Nat Protoc. 2014 Nov;9(11):2643–62.

21. Purcell S, Neale B, Todd-Brown K, Thomas L, Ferreira MAR, Bender D, et al. PLINK: A tool set for whole-genome association and population-based linkage analyses. Am J Hum Genet. 2007;81(3):559–75.

22. Jabbari E, Woodside J, Tan MMX, Shoai M, Pittman A, Ferrari R, et al. Variation at the TRIM11 locus modifies progressive supranuclear palsy phenotype. Ann Neurol [Internet]. 2018 [cited 2019 Sep 18];84(4):485–96.

23. Jabbari E, Koga S, Valentino RR, Reynolds RH, Ferrari R, Tan MMX, et al. Genetic determinants of survival in progressive supranuclear palsy: a genome-wide association study. 2021 Feb;20(2):107–116.

24. Kouri N, Ross OA, Dombroski B, Younkin CS, Serie DJ, Soto-Ortolaza A, et al. Genome-wide association study of corticobasal degeneration identifies risk variants shared with progressive supranuclear palsy. Nat Commun. 2015 Jun 16:6:7247.

25. Farrell K, Kim SH, Han N, Iida MA, Gonzalez EM, Otero-Garcia M, et al. Genome-wide association study and functional validation implicates JADE1 in tauopathy. Acta Neuropathol. 2022 Jan;143(1):33–53.

26. Ferrari R, Hernandez DG, Nalls MA, Rohrer JD, Ramasamy A, Kwok JBJ, et al. Frontotemporal dementia and its subtypes: A genome-wide association study. Lancet Neurol. 2014 Jul;13(7):686–99.

27. Watanabe K, Taskesen E, Van Bochoven A, Posthuma D. Functional mapping and annotation of genetic associations with FUMA. Nat Commun. 2017 Nov 28;8(1):1826.

28. Wang G, Sarkar A, Carbonetto P, Stephens M. A simple new approach to variable selection in regression, with application to genetic fine mapping. J R Stat Soc Series B Stat Methodol. 2020;82(5):1273–300.

29. Benner C, Spencer CCA, Havulinna AS, Salomaa V, Ripatti S, Pirinen M. FINEMAP: Efficient variable selection using summary data from genome-wide association studies. Bioinformatics. 2016;32(10):1493–501.

30. Andersson R, Gebhard C, Miguel-Escalada I, Hoof I, Bornholdt J, Boyd M, et al. An atlas of active enhancers across human cell types and tissues. Nature. 2014;507(7493):455–61.

31. Alexi N, Inge RH, Nicole GC, Johannes CMS, Miao Y, Rong H, et al. Brain cell type – specific enhancer – promoter interactome maps and disease-risk association. Science. 2019 Nov 29;366(6469):1134–1139.

32. Dunham I, Kundaje A, Aldred SF, Collins PJ, Davis CA, Doyle F, et al. An integrated encyclopedia of DNA elements in the human genome. Nature. 2012;489(7414):57–74.

33. Giambartolomei C, Vukcevic D, Schadt EE, Franke L, Hingorani AD, Wallace C, et al. Bayesian Test for Colocalisation between Pairs of Genetic Association Studies Using Summary Statistics. PLoS Genet. 2014;10(5).

34. Ardlie KG, DeLuca DS, Segrè A V., Sullivan TJ, Young TR, Gelfand ET, et al. The Genotype-Tissue Expression (GTEx) pilot analysis: Multitissue gene regulation in humans. Science (1979). 2015;348(6235):648–60.

35. Li YI, Knowles DA, Humphrey J, Barbeira AN, Dickinson SP, Im HK, et al. Annotation-free quantification of RNA splicing using LeafCutter. Nat Genet. 2018 Jan;50(1):151–158.

36. Parkinson TG, Program G. GP2: The Global Parkinson’s Genetics Program. Movement Disorders. 2021;36(4):842–51.

37. Ren Y, Liu Y, Zheng S, Luo M. KCTD8 and KCTD12 Facilitate Axonal Expression of GABA B Receptors in Habenula Cholinergic Neurons. The Journal of Neuroscience. 2022;42(9):1648–65.

38. Zheng S, Abreu N, Levitz J, Kruse AC. Structural basis for KCTD-mediated rapid desensitization of GABAB signalling. Nature. 2019 Mar;567(7746):127–131.

39. Zhu Y, Afolabi LO, Wan X, Shim JS, Chen L. TRIM family proteins: roles in proteostasis and neurodegenerative diseases. Open Biol. 2022;12(8).

40. Kang C, Lu Z, Zhu G, Chen Y, Wu Y. Knockdown of TRIM22 Relieves Oxygen-Glucose Deprivation/Reoxygenation-Induced Apoptosis and Inflammation Through Inhibition of NF-κB/NLRP3 Axis. Cell Mol Neurobiol. 2021 Mar;41(2):341–351.

41. Forlani G, Accolla RS. Tripartite motif 22 and class II transactivator restriction factors: Unveiling their concerted action against retroviruses. Front Immunol. 2017;8(OCT):1–7.

42. Ozato K, Shin DM, Chang TH, Morse HC. TRIM family proteins and their emerging roles in innate immunity [Internet]. Nat Rev Immunol. 2008 Nov;8(11):849–60.

43. Dhaenens CM, Tran H, Frandemiche ML, Carpentier C, Schraen-Maschke S, Sistiaga A, et al. Mis-splicing of Tau exon 10 in myotonic dystrophy type 1 is reproduced by overexpression of CELF2 but not by MBNL1 silencing. Biochim Biophys Acta Mol Basis Dis. 2011 Jul;18127):732–42.

44. Metz M, Gassmann M, Fakler B, Schaeren-Wiemers N, Bettler B. Distribution of the auxiliary GABAB receptor subunits KCTD8, 12, 12b, and 16 in the mouse brain. Journal of Comparative Neurology. 2011 Jun 1;519(8):1435–54.

45. Bocchetta M, Gordon E, Marshall CR, Slattery CF, Cardoso MJ, Cash DM, et al. The habenula: An under-recognised area of importance in frontotemporal dementia? J Neurol Neurosurg Psychiatry. 2016;87(8):910–2.

46. Bocchetta M, Malpetti M, Todd EG, Rowe JB, Rohrer JD. Looking beneath the surface: the importance of subcortical structures in frontotemporal dementia. Brain Commun. 2021;3(3).

47. Hof PR, Bouras C, Perl DP, Morisson JH. Acta Neuropathol. Quantitative neuropathologic analysis of Pick’s disease cases: cortical distribution of Pick bodies and coexistence with Alzheimer’s disease1994;87(2):115–24.

48. Alazami AM, Patel N, Shamseldin HE, Anazi S, Al-Dosari MS, Alzahrani F, et al. Accelerating novel candidate gene discovery in neurogenetic disorders via whole-exome sequencing of prescreened multiplex consanguineous families. Cell Rep. 2015;10(2):148–61.

49. Faqeih EA, Almannai M, Saleh MM, AlWadei AH, Samman MM, Alkuraya FS. Phenotypic characterization of KCTD3-related developmental epileptic encephalopathy. Clin Genet. 2018;93(5):1081–6.

50. Michaelson JJ, Shi Y, Gujral M, Zheng H, Malhotra D, Jin X, et al. Whole-genome sequencing in autism identifies hot spots for de novo germline mutation. Cell. 2012;151(7):1431–42.

51. Weiss L. Association between Microdeletion and Microduplication at 16p11.2 and Autism. N Engl J Med. 2008 Feb 14;358(7):667–75.

52. McCarthy SE, Makarov V, Kirov G, Addington AM, McClellan J, Yoon S, et al. Microduplications of 16p11.2 are associated with schizophrenia. Nat Genet. 2009;41(11):1223–7.

53. Mencacci NE, Rubio-Agusti I, Zdebik A, Asmus F, Ludtmann MHR, Ryten M, et al. A Missense Mutation in KCTD17 Causes Autosomal Dominant Myoclonus-Dystonia. Am J Hum Genet. 2015;96(6):938–47.

54. Graziola F, Stregapede F, Travaglini L, Garone G, Verardo M, Bosco L, et al. A novel KCTD17 mutation is associated with childhood early-onset hyperkinetic movement disorder. Parkinsonism Relat Disord. 2019 Apr:61:4–6.

55. Marcé-Grau A, Correa M, Vanegas MI, Muñoz-Ruiz T, Ferrer-Aparicio S, Baide H, et al. Childhood onset progressive myoclonic dystonia due to a de novo KCTD17 splicing mutation. Parkinsonism Relat Disord. 2019;61(October 2018):7–9.

56. Lee MTM, Chen CH, Lee CS, Chen CC, Chong MY, Ouyang WC, et al. Genome-wide association study of bipolar i disorder in the Han Chinese population. Mol Psychiatry. 2011;16(5):548–56.

57. Metz KA, Teng X, Coppens I, Lamb HM, Wagner BE, Rosenfeld JA, et al. KCTD7 deficiency defines a distinct neurodegenerative disorder with a conserved autophagy-lysosome defect. Ann Neurol. 2018;84(5):766–80.

58. Wang Y, Cao X, Liu P, Zeng W, Peng R, Shi Q, et al. KCTD7 mutations impair the trafficking of lysosomal enzymes through CLN5 accumulation to cause neuronal ceroid lipofuscinoses. Sci Adv. 2022;8(31):1–16.

59. Ferrer I. Neurons and their dendrites in frontotemporal dementia. Dement Geriatr Cogn Disord. 1999:10 Suppl 1:55–60

60. Leuzy A, Zimmer ER, Dubois J, Pruessner J, Cooperman C, Soucy JP, et al. In vivo characterization of metabotropic glutamate receptor type 5 abnormalities in behavioral variant FTD. Brain Struct Funct. 2016;221(3):1387–402.

61. Murley AG, Rouse MA, Simon Jones P, Ye R, Hezemans FH, O’Callaghan C, et al. GABA and glutamate deficits from frontotemporal lobar degeneration are associated with disinhibition. Brain. 2021;143(11):3449–62.

62. Mandell MA, Jain A, Arko-Mensah J, Chauhan S, Kimura T, Dinkins C, et al. TRIM Proteins Regulate Autophagy and Can Target Autophagic Substrates by Direct Recognition. Dev Cell. 2014;30(4):394–409.

63. Kimura T, Mandell M, Deretic V. Precision autophagy directed by receptor regulators - emerging examples within the TRIM family. Vol. 129, Journal of Cell Science. 2016. 881–891 p.

64. Chen L, Zhu G, Johns EM, Yang X. TRIM11 activates the proteasome and promotes overall protein degradation by regulating USP14. Nat Commun. 2018 Dec 1;9(1).

65. Zhang L, Afolabi LO, Wan X, Li Y, Chen L. Emerging Roles of Tripartite Motif-Containing Family Proteins (TRIMs) in Eliminating Misfolded Proteins. Front Cell Dev Biol. 2020 Aug 25;8:802.

66. Giraldo MI, Hage A, van Tol S, Rajsbaum R. TRIM Proteins in Host Defense and Viral Pathogenesis. Curr Clin Microbiol Rep. 2020;7(4):101–114.

67. Deng NH, Zhou ZX, Liu HT, Tian Z, Wu ZF, Liu XY, et al. TRIMs: Generalists Regulating the NLRP3 Inflammasome Signaling Pathway. DNA Cell Biol. 2022;41(3):262–75.

68. Mishra A, Duplaà C, Vojinovic D, Suzuki H, Sargurupremraj M, Zilhão NR, et al. Gene-mapping study of extremes of cerebral small vessel disease reveals TRIM47 as a strong candidate. Brain. 2022;145(6):1992–2007.

69. Johnson K, De Ridder W, Töpf A, Bertoli M, Phillips L, De Jonghe P, et al. Extending the clinical and mutational spectrum of TRIM32 -related myopathies in a non-Hutterite population. J Neurol Neurosurg Psychiatry. 2019;90(4):490–3.

70. Tissot C, Mechti N. Molecular cloning of a new interferon-induced factor that represses human immunodeficiency virus type I long terminal repeat expression. Journal of Biological Chemistry. 1995;270(25):14891–8.

71. Chen L, Brewer MD, Guo L, Wang R, Jiang P, Yang X. Enhanced Degradation of Misfolded Proteins Promotes Tumorigenesis. Cell Rep. 2017;18(13):3143–54.

72. Janer A, Martin E, Muriel MP, Latouche M, Fujigasaki H, Ruberg M, et al. PML clastosomes prevent nuclear accumulation of mutant ataxin-7 and other polyglutamine proteins. Journal of Cell Biology. 2006;174(1):65–76.

73. Forlani G, Tosi G, Turrini F, Poli G, Vicenzi E, Accolla RS. Tripartite motif-containing protein 22 interacts with class II transactivator and orchestrates its recruitment in nuclear bodies containing TRIM19/PML and Cyclin T1. Front Immunol. 2017 May 15:8:564.

