## Supplementary Materials for "Identification of common variants influencing risk of the three-repeat tauopathy Pick’s disease: a genome wide association study"

#### **(A) Supplementary Methods and Results – Main Analysis**

#### **Supplementary Methods**

**DNA Extraction**

For the majority of cases, DNA was extracted at their respective collection site (MCJ or UCL). At UCL genomic DNA was extracted from either frozen brain tissue or whole blood lymphocytes using the Kleargene XL Nucleic Acid Purification kit (LGC, Germany). At MCJ genomic DNA was extracted from frozen brain tissue from PiD cases and from peripheral blood lymphocytes from control subjects using an automated or manual method. Automated DNA extractions were carried out using Autogen Tissue Kit reagents according to manufacturer protocols and were processed on the Autogen FlexSTAR+ (both Autogen, Holliston, MA, USA). Manual extractions were completed using QIAamp DNA Mini Kits (Qiagen, MD, USA). DNA quality for all samples was assessed with a NanoDrop 8000 spectrophotometer (ThermoFisher Scientific, USA) and absorbance ratios for 260/280 and 260/230 were between 1.7-2.2 and 2.0-2.2, respectively. Sydney Brain Bank extracted DNA from peripheral blood using a Qiagen DNA extraction kit, UPenn and UCSF extracted DNA from their cases using QIAamp DNA mini kits.

**Quality Control and Imputation**

Samples with a low overall genotyping rate (<0.98), related individuals (Identity-By-Descent PIHAT>0.1), and heterozygosity outliers (>2SDs away from the mean) were removed, as were individuals where clinically reported biological sex did not match genetically determined sex. We excluded variants with a missingness rate > 0,05, minor allele frequency (MAF) < 0.01 and Hardy-Weinberg equilibrium (HWE) $p$ < 1 × 10^-6^. Principal component analysis (PCA) was performed on a linkage disequilibrium (LD) pruned set of variants (removing SNPs with an r^2^ > 0.5 in a 50kb sliding window shifting five SNPs at a time), after merging with the HapMap3 reference panel (**Supp. Figure 9A** and **Supp. Figure 9B**) for NBA and GSA datasets respectively). Individuals who deviated more than six standard deviations (6SD) from the mean of the first 10 principal components of the HapMap3 CEU population were excluded from the analysis.

The two genotyping array datasets (NBA and GSA) were imputed separately against the Haplotype Reference Consortium (HRC) reference panel (version r1.1 2016; http://www.haplotype-reference-consortium.org/) in the Michigan Imputation Server (RRID:SCR_017579; https://imputationserver.sph.umich.edu) using Minimac4 (version 1.0.0; https://genome.sph.umich.edu/wiki/Minimac4). Imputed variants were excluded if the imputation information R^2^ was $\leq$ 0.7 and the genotype posterior probability was $\leq$ 0.9, to ensure that only high-quality genotype calls were retained for further analysis. We then merged the two datasets based on shared variants, and variants with missingness >5% and minor allele frequencies < 1% were also excluded. Ancestry was then rechecked on the merged dataset using the same procedure as detailed above, excluding any further samples that were greater than 6SD from the mean of the first 10 principal components of the CEU population (**Supp. Figure 9C**). After this final extraction of European-ancestry samples the first ten principal components were re-calculated and used as these as covariates in the association analysis

**Genomic risk loci definition and gene mapping**

The settings used in Functional Mapping and Annotation of Genome-Wide Association Studies (FUMA)were as follows. Genomic risk loci within 250 kilobases (kb) of each other were incorporated into the same locus. The individual genomic risk loci were mapped to genes using positional and eQTL mapping. For positional mapping all variants within 10kb of a gene in the genomic risk locus were assigned to that gene. For eQTL mapping, FUMA maps variants based on significant eQTL interactions in the PsychEncode, BloodeQTL, CMC, and GTEXv8 (Brain and whole blood) data repositories. GTEXv8 was used to identify whether any of the lead SNPs were eQTLs for gene expression. Regional association plots were generated in LocusZoom (RRID:SCR_021374; <https://my.locuszoom.org/>), and LDProxy was used to identify any deleterious variants in high linkage disequilibrium (LD) with variants of interest (https://ldlink.nci.nih.gov/?tab=ldproxy)(1) in European populations (excluding the Finnish population).

**Conditional analysis**

To understand whether there were one or more variants at the same locus contributing to the signal at each genomic risk locus, we performed a conditional analysis for the lead 5 loci conditioning on the lead SNP at that loci.(2) We used the GWAS summary statistics and the AMP-PD cohort (*n* = 10,418) as the reference sample for linkage disequilibrium estimation. The reference sample went through the same QC steps as described above for the PiD cohort. We then used CGTA-COJO (v.1.93.0 beta for Linux; <https://yanglab.westlake.edu.cn/software/gcta/#Overview>)(3) to perform association analyses conditioned on SNPs of interest.

**Fine-mapping and functional annotation**

A credible set, in the context of a multiple regression model, is defined as a subset of variants that has a 95% probability of $p$, or greater, of containing at least one effect variant (i.e. a variant with a non-zero regression coefficient). SuSiE calculates the credible set using a simple model-fitting algorithm, Iterative Bayesian Stepwise Selection (IBSS), that fits a “single effects" regression model at each step(4). FINEMAP on the other hand uses a Shotgun Stochastic Search (SSS) algorithm that explores the causal configuration space by concentrating effort on those configurations with non-negligible probability(5). To compute the SNPs correlation matrix necessary to infer the SNPs credible set, we used the 503 European ancestry individuals from the Phase 3 of the 1000 Genomes Project.

**Colocalisation analysis**

We used cortex-specific cis-eQTLs from MetaBrain (https://www.metabrain.nl/)(6). Coloc was run using default priors; these are the prior probabilities that any random SNP in the region is associated with trait 1 or trait 2, $p$1=1 × 10^-4^ and $p$2=1 × 10^-4^. A threshold of $p$12=5 × 10^-6^ was used for the $p$12 prior, which is the probability that a SNP in the region is associated with both traits. Loci with a posterior probability of hypothesis 4 (PP.H4) ≥ 0.85 were considered as significant evidence of colocalization between the GWAS and the eQTL traits (one shared causal variant). We used a more conservative threshold for PP.H4 than used in the original work by Giambartolomei et al(7) (0.75), in line with more recent work where a threshold of 0.8593 or even 0.9085 has been used on GWAS data, to try and minimise the false positive rate.

**Assessment of gene transcript and protein expression of lead genes**

Brain expression profiles of gene transcripts and encoded proteins highlighted by the GWAS were assessed, using a range of different publicly available online data sources. Bulk RNA and protein expression was assessed using The Human Protein Atlas (RRID:SCR_006710; <https://www.proteinatlas.org/>)(8). This resource provides consensus RNA expression data derived from the GTEXv8 RNA-seq, Human Protein Atlas (HPA) RNA-seq and the FANTOM CAGE datasets. Protein expression by tissue is based on tissue profiles generated from 6120 antibodies with more than five million immunohistochemistry-based images covering 5067 human genes, corresponding to approximately 25% of the human genome(9).

Cell specific RNA expression was investigated using the Brain RNA-Seq database (RRID:SCR_017483; <https://www.brainrnaseq.org/>)(10), and single cell RNA-seq data provided by DropViz (<http://dropviz.org/>)(11). Data in the Brain RNA-seq dataset was generated from healthy temporal lobe samples resected from 14 patients intra-operatively. The cell types sequenced from these samples were mature astrocytes (n=12), microglia (n=3), oligodendrocytes (n=3) and neuron (n=1). DropViz provides gene expression on 690,000 individual cells derived from nine different regions of the adult mouse brain. Predicted protein interaction networks were investigated using the STRING database (RRID:SCR_005223; <https://string-db.org/>).

#### **Supplementary Results**

***MAPT* mutations with 3R tau pathology**

All samples included from MCJ and Sydney had negative *MAPT* mutation screening. All UCL samples were screened for known *MAPT* mutations covered by the NBA at the GenomeStudio genotype calling stage. This identified four PiD cases that had a total of five rare *MAPT* variants; one case had K280del (GRCh37/hg19 Chr17:44087694 AAG>---), the second case had A239T (Chr17:44073923 G>A; p.Ala239Thr), the third case had both S318L (Chr17:44061123 C>T, p.Ser319Leu) and V363I (Chr17:44096073 G>A; p.Val363Ile), and the fourth case had Q230R (Chr17:44060859 A>G; p.Gln305Arg). Of these four PiD cases, two of them failed QC (A239T IBD, S318L and V363I > 3SD HZ) so were excluded from the association analysis. The K280del was excluded from downstream analysis as it is likely to be pathogenic, while the Q230R was included given this is likely a benign polymorphism in *MAPT* with a MAF of 0.05 in the European (non-Finish) population (GnomAD; https://gnomad.broadinstitute.org/). The MAF for this variant tagging Q230R in our dataset was 0.06 for both cases and controls, supporting the fact that this is unlikely to be a deleterious variant in PiD.

***MAPT* Haplotype Frequency**

Analysis of *MAPT* H1 and H2 haplotype frequency showed an increase (Chi square: χ = 6.04, df =2, p = 0.003) in both H1/H2 heterozygotes (45.6% PiD cases vs. 36.1% controls) and H2/H2 homozygotes (6.8% PiD cases vs. 5% controls) (**Supp. Table 3**).

**Transcript and protein expression of suggestive genes**

*KCTD8* transcript expression is enriched in the central nervous system (**Supp. Figure 7A**); the cerebellum has the highest expression levels (normalised transcripts per million (nTPM) 16), with expression in the cerebral cortex, amygdala and hippocampus at 7.0, 6.4 and 5.4 nTPM respectively. Human single cell RNA-seq data shows that oligodendroglia are the most enriched cell type (576.7 nTPM) followed by inhibitory neurons (435.3 nTPM), then excitatory neurons (219.9 nTPM). Astrocytes and microglia are the least enriched of the brain cell types (164.2 and 69.3 nTPM respectively) (**Supp. Figure 7B**). Mouse scRNA data suggests that habenula neurons demonstrate high and specific KCTD8 RNA expression (**Supp. Figure 7C**). At the protein level, in keeping with RNA expression levels, immunohistochemistry demonstrates that *KCTD8* is most highly expressed in the cerebellum and the cerebral cortex (**Supp. Figure 7D**). **Supp. Figure 7E** shows the predicted protein interaction network for KCTD8 protein, highlighting its interactions with GABRG1, GABRA4 and GABRB1 all of which are subunits of the GABA_a_ receptor in the human brain.

The *TRIM22* gene on the other hand has low tissue specificity and is ubiquitously expressed throughout the body, with highest expression in lymphoid tissues (spleen nTPM 143.1) (**Supp. Figure 8A**). *TRIM22* however, still expressed in the brain with transcript expression in the brain ranging from 12.8 nTPM in the thalamus to 29.3 nTPM in the white matter. This low specificity is also reflected at the level of protein expression, with levels in the brain ranging from low in the cerebellum and hippocampus to medium in the cerebral cortex and caudate (**Supp. Figure 8B**). The Brain RNA-seq data shows that *TRIM22* is predominantly expressed microglia within the brain (FPKM; 39.07 $\pm$ 3.36) , with little expression in neurons (FPKM; 0.632) (**Supp. Figure 8C**). Unfortunately, *TRIM22* is not included in the mouse scRNA dataset so it was not possible to interrogate its single cell RNA specificity. **Supp. Figure 8D** shows the predicted protein interaction network for *TRIM22*. Of interest is the interaction with promyelocytic leukaemia protein (*PML* also known as *TRIM19)* a protein that promotes clearance of misfolded proteins (including mutant ataxin-7 in Spinal Cerebellar Ataxia Type 7)(12) via the ubiquitin proteosome system (UPS)(13). *PML* has been shown to colocalise with *TRIM22* in nuclear complexes upon IFN-γ induced *TRIM22* expression(14).

Although *YIPF7* is predominantly expressed in skeletal muscle (nTPM 84.9), with low transcript levels in the bulk brain tissue (cerebellum 5.4 nTPM, cerebral cortex 1.3 nTPM) (**Supp. Figure 9A**), interestingly antibody staining in the Human Protein Atlas suggests that the protein is present in the brain with high levels detected in glial cells in the hippocampus and medium levels in neurons of the cerebral cortex, caudate and cerebellum (**Supp. Figure 9B**). In addition, although overall *YIPF7* RNA expression is low in the brain, its expression is enriched specifically within excitatory and inhibitory neurons (**Supp. Figure 9C**). Further investigation of Y*IPF7*’s protein interaction network yields some interesting findings (**Supp. Figure 9D**). Firstly, *YIPF7* is predicted to interact with both *YIF1A* and *YIF1B* to form the YIPF complex 1(15), that is localised to the early compartment between the endoplasmic reticulum (ER) and the Golgi, and participates in the anterograde recycling of proteins from the ER to the cell membrane. *YIF1A* has been shown to bind *VAPB*; the *VAPB* P56S mutation is known to cause amyotrophic lateral sclerosis(16). Homozygous mutations in *YIF1B* have recently been shown to cause a progressive encephalopathy with global developmental delay and cognitive impairment(17,18). Secondly, Y*IPF7* is also predicted to interact with *dystrophica myotonica-protein-kinase* (*DMPK*), mutations in which cause myotonica dystrophy type 1 (DM1). This is of particular interest given the known association between DM1 and predominantly 3R-tau pathology in the brain at post-mortem(19,20)

#### **(B) Supplementary Methods and Results: Batch Effect Characterization and Sensitivity Analyses**

#### **Overview**

To ensure that our genome‑wide association study (GWAS) results are robust and not confounded by technical or cohort‑specific effects, we performed comprehensive batch effect characterization and sensitivity analyses. Herein we detail characterization of systematic differences between genotyping platforms, additional analytical approaches to address potential confounding, and compare results across multiple modelling strategies.

#### **1. Characterization of Batch Effects**

**1.1 Covariate Selection**

Given this was the first GWAS for the rare disease PiD with a small cohort where confounders were unknown, an unbiased, data-driven stepwise regression was used to identify the optimal statistical model. For comparison an additional model was run adjusting for gender, genotype array, age and the first three genetic principal components (PCs 1-3). A comprehensive comparison is included in section 2.

**1.2 Control‑Control GWAS: Quantifying Genetic Batch Effects**

To systematically characterize genetic differences between genotyping platforms, we performed a control‑control GWAS treating genotyping array (NBA vs. GSA) as the phenotype.

Methods: Logistic regression was performed on controls only, testing association between each SNP and genotyping platform:

$logit\left( P\left( Array=GSA \right) \right)= \beta_{0}+ \beta_{1}\left( AGE \right)+ \beta_{2}\left( SEX \right)+ \beta_{3}\left( PC1 \right)+ \ldots+ \beta_{11}\left( PC10 \right)+ \beta_{SNP}\left( SNP \right)$

Where AGE = Age at death (cases) or age at blood draw (controls), SEX = biological sex (0 = male, 1 = female), PC1‑10 = first ten principal components from ancestry analysis, and SNP = additive genetic model (0, 1, or 2 copies of effect allele). The first 10 principal components were examined for association with genotyping array (**Supp Table 6**). Multiple principal components showed significant association with genotyping array(Array=1=GSA) (PC1, PC2, PC3, PC5, PC7, PC10: all P < 0.01) and Age was strongly associated with array membership (p < 0.001), confirming the presence of systematic genetic and demographic differences between platforms. Our analysis indicated that most of the variance between genotyping arrays is captured by PC1 (**Supp. Figure 10**).

#### **2. Confounding and Covariate Sensitivity**

To ensure our findings are robust to potential confounding by genotyping platform and demographic differences, we implemented three complementary analytical strategies:

**2.1 Modified Primary Model:**

Standard approach for multi cohort GWAS with greater power to detect medium sized effects. PC1, 2, and 3 selected instead of those identified through step regression as below.

$$logit\left( P\left( PiD=1 \right) \right)= \beta_{0}+ \beta_{1}\left( ARRAY \right)+ \beta_{2}\left( AGE \right)+ \beta_{3}\left( SEX \right)+ \beta_{4}\left( PC1 \right)+ \beta_{5}\left( PC2 \right)+ \beta_{6}\left( PC3 \right)+ \beta_{SNP}\left( SNP \right)$$

Where:

ARRAY = genotyping platform (0 = NBA, 1 = GSA)

Resulting in 294 cases, 1,055 controls with 6,316,457 SNPs passing quality control. The Genomic Inflation Factor λ = 0.99 (λ₁₀₀₀ = 0.97)

**2.2 Model 2: Interaction Model**

Given the substantial age difference between GSA cases and controls (14.4 years), we tested whether the age effect differs by genotyping platform. Genome‑wide analysis was performed in parallel across 500 genomic partitions with each SNP tested with the full interaction model.

$$logit\left( P\left( PiD=1 \right) \right)= \beta_{0}+ \beta_{1}\left( ARRAY \right)+ \beta_{2}\left( AGE \right)+ \beta_{3}\left( SEX \right)+ \beta_{4}\left( PC1 \right)+ \beta_{5}\left( PC2 \right)+ \beta_{6}\left( PC3 \right)+ \beta_{7}\left( ARRAY\times AGE \right)+ {\beta_{SNP}\left( SNP \right)}$$

The interaction term *β₇(ARRAY × AGE)* allows the effect of age on PiD risk to vary by genotyping platform, accounting for the different age distributions between NBA and GSA cohorts. This approach explicitly models the non‑constant age effect across platforms, producing more robust effect size estimates when age distributions are non‑overlapping.

**2.3 Model 3: Array‑Specific GWAS and Meta‑Analysis**

To assess whether associations are driven by a single cohort or are consistent across both platforms, we performed separate GWAS for each array followed by fixed‑effects meta‑analysis. Details on methods for meta-analysis are given in **Supplementary Methods A**.

**Step 1 ‑ Array‑Specific GWAS**:

GSA‑only Analysis (159 cases, 75 controls) and NBA‑only Analysis (135 cases, 980 controls):

$$logit\left( P\left( PiD=1 \right) \right)= \beta_{0}+ \beta_{1} \left( AGE \right)+ \beta_{2} \left( SEX \right)+ \beta_{3} \left( PC1 \right)+ \beta_{4} \left( PC2 \right)+ \beta_{5} \left( PC3 \right)+ {\beta_{SNP}\left( SNP \right)}$$

**Step 2 ‑ Meta‑Analysis**:

Fixed‑effects inverse variance‑weighted meta‑analysis using *metal* software:

$$\beta\_meta=((\beta\_GSA\times w\_GSA+ \beta\_NBA\times w\_NBA ))/ (w\_GSA+ w\_NBA )$$

where w_i = 1 / SE_i²

The array-stratified GWAS meta-analysis did not identify any disease-associated variants reaching genome-wide significance ($p$< 5 × 10^-8^), but two loci showed a suggestive association (**Supp. Table 7**) (defined as defined as $p$ < 5 × 10^-6^); rs72651516 (OR = 2.94, 95% CI = 1.90-4.55, $p$ = 1.83 × 10^-6^) an intergenic SNP (nearest gene *HMGN2*) and rs117040196 (OR = 2.94, 95% CI = 1.90-4.55, $p$ = 1.83 × 10^-6^) an intronic SNP located in *SYNDIG1* (OR 0.20, 95% CI = 0.10-0.39, $p$ = 4.57 × 10^-6^) (**Supp. Figure 4** – lower half of mirror Manhattan plot). **Supp. Figure 11** shows more detailed regional association plots at these two loci. The genomic inflation factor ($\lambda$) was 0.98 ($\lambda_{1000}$ = 0.96) confirming no population stratification (**Supp Figure 2)**

rs72651516 is an eQTL for LOC284798, an uncharacterised ncRNA predominantly expressed in the testis and the nucleus accumbens in the brain. rs117040196 is an eQTL for *DDHS*, a gene upstream of *HMGN2*, responsible for producing an enzyme that synthesises dolichol, a lipid essential for N-linked glycosylation. Heterozygous mutations in this gene have been associated with a fatal Type I congenital disorder of glycosylation(21), developmental delay and seizures with or without movement abnormalities(22), while homozygous mutations have been associated with non-syndromic retinitis pigmentosa (RP type 59)(23).

#### **3. Comparison of Results Across Analytical Approaches**

**3.1 Top Associated Loci from Primary Analysis**

Five loci reached suggestive genome‑wide significance ($p$ < 5 × 10⁻⁶) in the primary combined analysis (Table 3- main manuscript). We compared results for these loci across all three alternative analytical approaches.

**3.2 Interpretation of Results Across Models**

**Our analysis shows robust association for 4 out of five hits identified by the combined method.**

**KCTD8 (4:44392571)**: All three approaches show consistent association ($p$ < 10⁻⁵) and effect sizes are comparable across models (OR: 5.3‑7.53). Signal primarily driven by larger NBA cohort, the GSA‑only analysis is underpowered as evident by the very large SE, however, direction remains consistent. Our results are indicative of robust association, not solely driven by batch effects.

**TRIM22 (11:5724803):** Consistent association across simple combined and interaction models ($p$ < 10⁻⁵). Meta‑analysis shows suggestive association ($p$ = 1.3 × 10⁻⁴) possibly due to loss of power. Interestingly the lead SNP rs7936441 (OR 1.94, 95% CI 1.40 – 2.69, ($p$ = 6.47 x 10^-5^) at Chromosome 11 (TRIM22) locus in the meta-analysis is in complete linkage disequilibrium with rs66481907. Effect sizes are stable (OR: 1.9‑2.4).

**RANBP3L (5:36376351) and GABRG3 (15:27729149)**: Also consistent in both the combined approaches (simple and interaction) as well as the meta-analysed data.

**RYR1 (19:39029201)**: Inconsistent directions of effect between GSA and NBA, with GSA‑only analyses showing unstable estimates (wide CIs, opposite directions). The meta‑analysis reveals substantial heterogeneity (Direction: ‑+) indicating that the hit is likely driven by batch effects or population stratification.

**(C) Supplementary Figures**

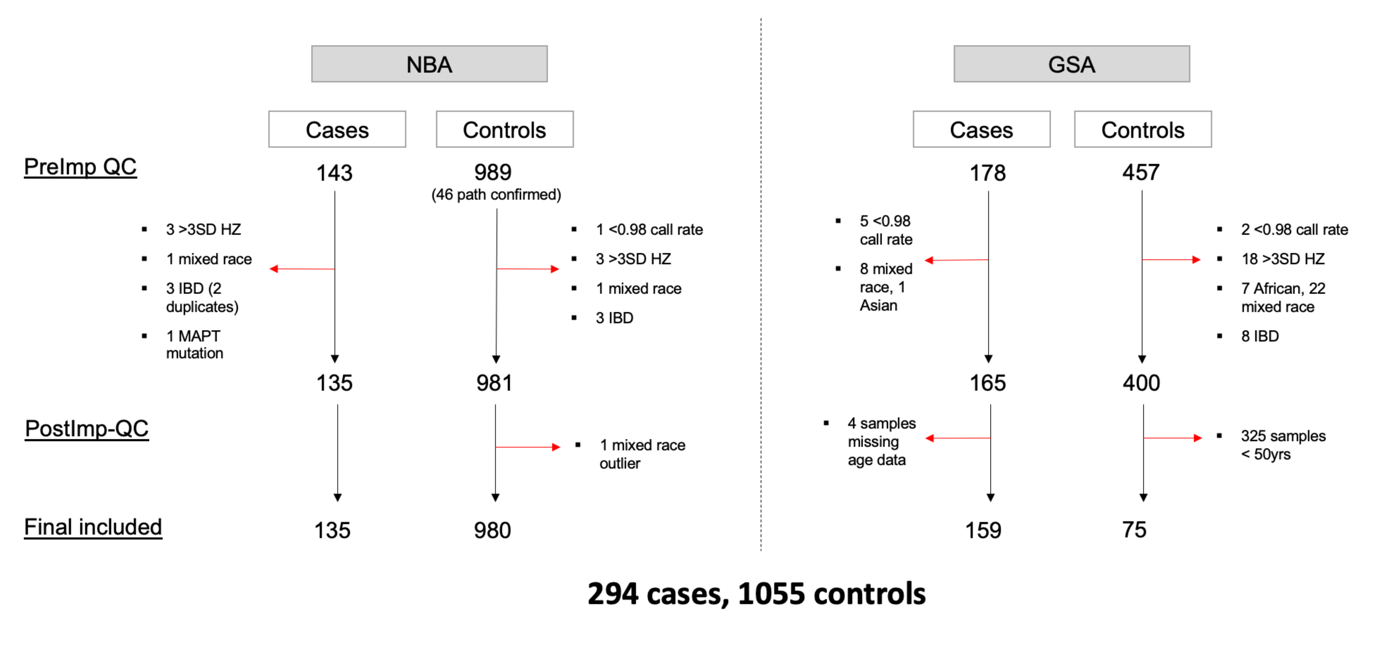

**Supplementary Figure 1 - Overview of sample quality control for PiD GWAS.** Summary of samples excluded at each stage of quality control (QC) pre-imputation (PreImp) and post-imputation (PostImp). Final number of samples that passed QC included in GWAS detailed at bottom of figure. NBA = Illumina NeuroBooster Array, GSA = Illumina Global Screening Array

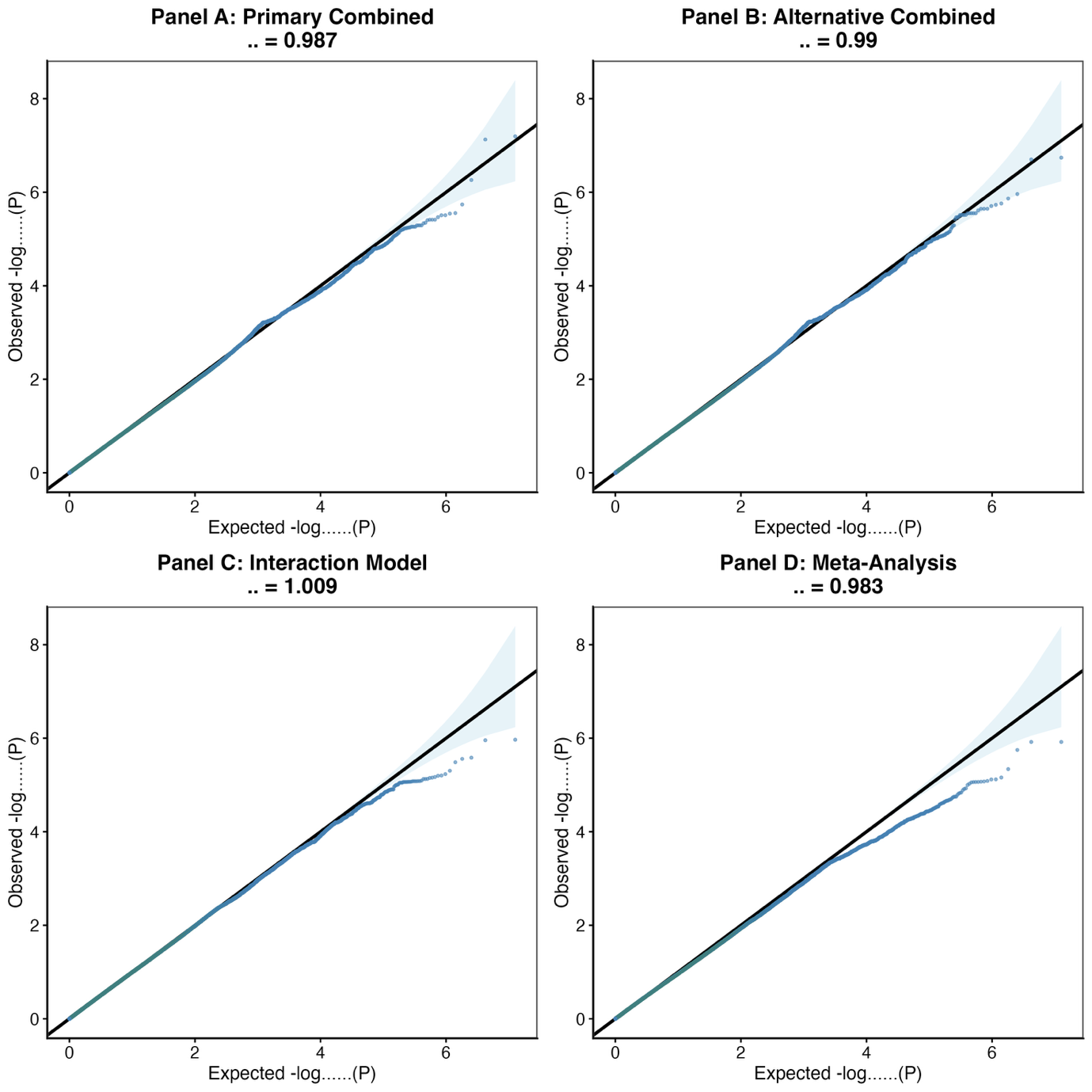

**Supplementary Figure 2 – Quantile-quantile (QQ) plots for different GWAS models** **(A)** Primary combined model adjusted for age, gender, genotype array and PC1, 8 and 10 **(B)** Alternative combined model adjusted for age, gender, genotype array and PC1-3 **(C) Interaction model** with addition of an Array $\times$ Age term in addition to covariates included in the alternative combined model **(D)** Meta-analysis model adjusted for age, gender, and PC1-3. Lambda = genomic inflation factor ($\lambda$).

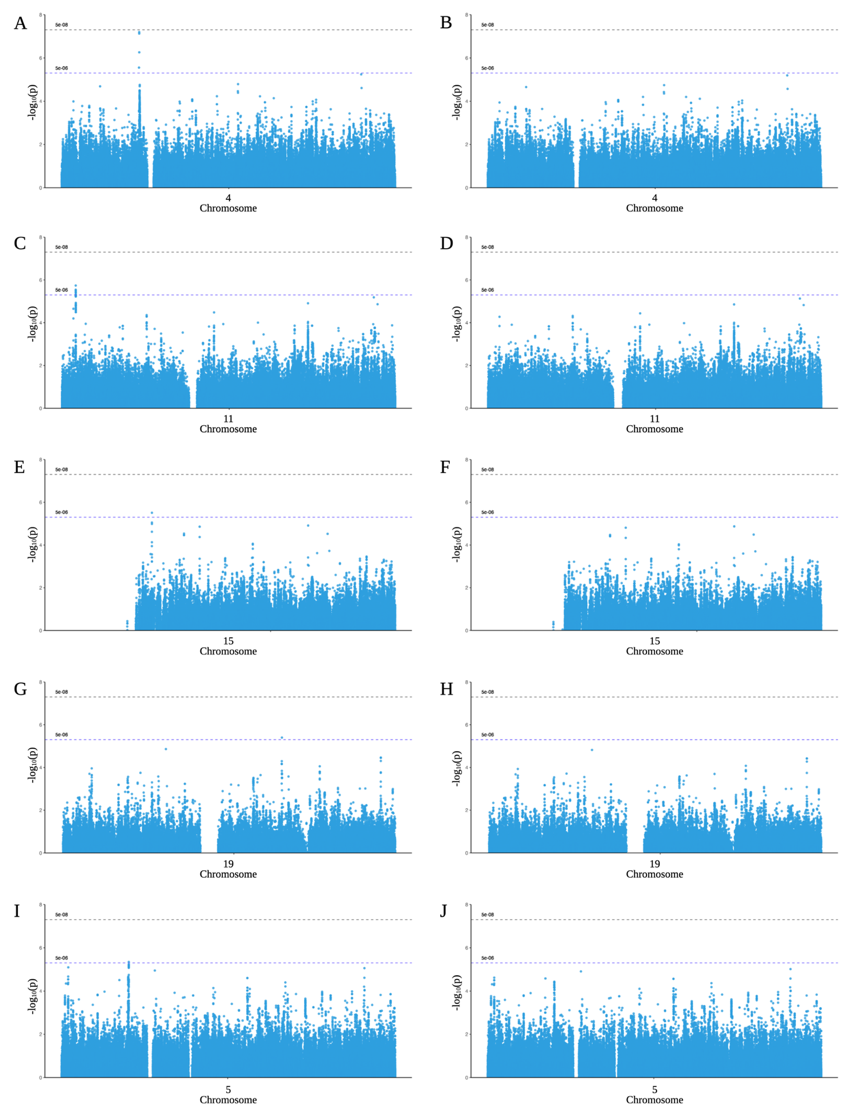

**Supplementary Figure 3 - Conditional analyses adjusted for the lead SNP at each suggestive GWAS loci.** For chromosome 4 (**A**) Unconditioned Manhattan plot (**B**) Manhattan plot conditioned on the *KCTD8* variant rs112161979. For chromosome 11 (**C**) Unconditioned Manhattan plot and **(D**) Manhattan plot conditioned on the *TRIM22* variant rs66481907. For chromosome 15 **(E**) Unconditioned Manhattan plot and (**F**) Manhattan plot conditioned on the *GABRG3* variant rs112721576. For chromosome 19 (**G**) Unconditioned Manhattan plot and (**H**) Manhattan plot conditioned on the *RYR1* variant rs11881082. For chromosome 5 (**I**) Unconditioned Manhattan plot and (**J**) Manhattan plot conditioned on the intergenic variant rs7720520 near *RANBP3L*.

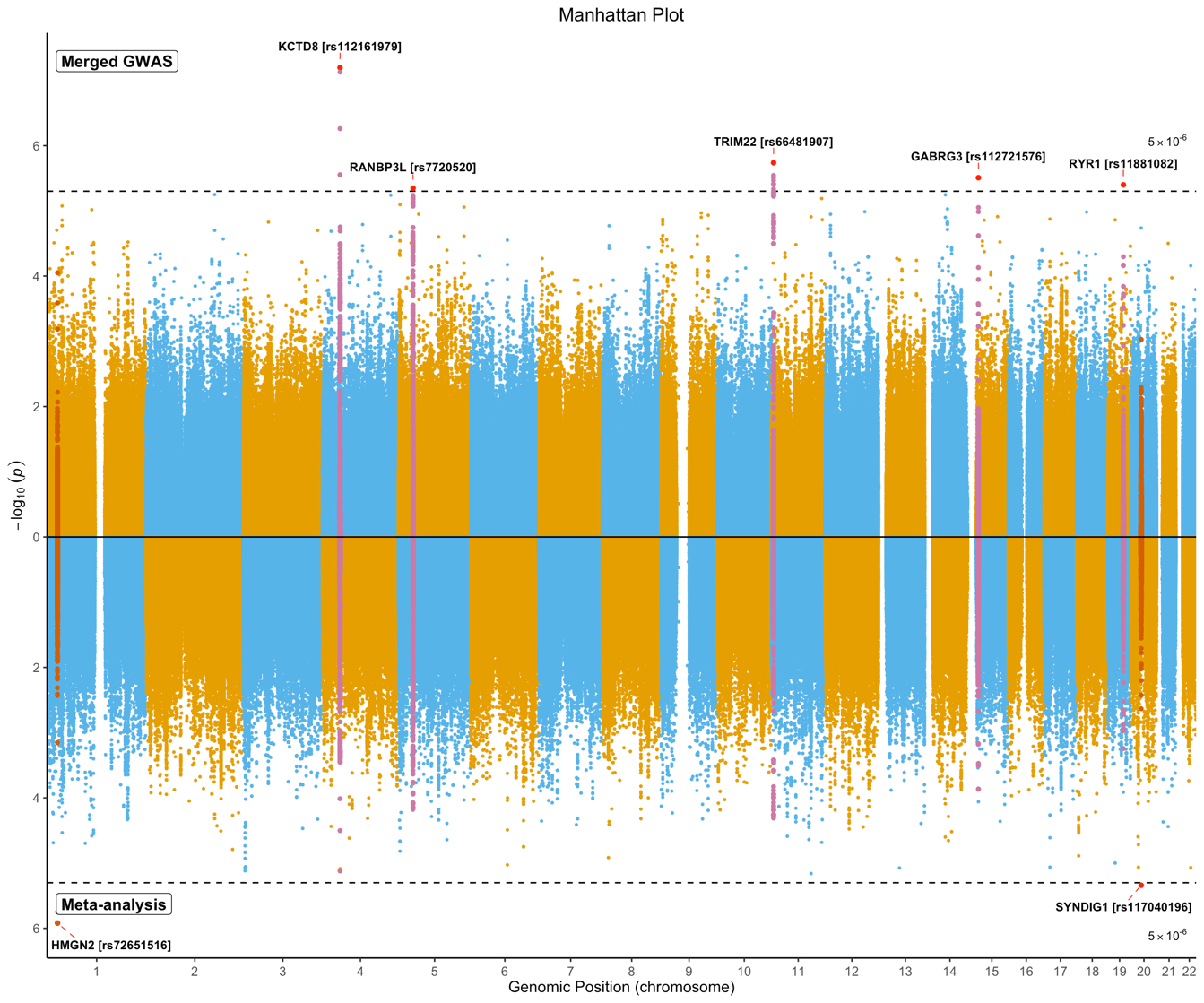

**Supplementary Figure 4: Mirror Manhattan plot** comparing -log10$(p$) values from logistic regression of imputed variants corrected for age, gender, array and three principal components (PC1, 8 and 10) on the top of the plot (Merged GWAS), and log10 $(p$) values from the GWAS metanalysis using summary statistics for the cohorts genotyped on NBA and GSA via logistic regression adjusted for standardised age, sex, and three PCs (meta-analysis). Red dots indicate the variant (rsID and nearest gene labelled) with the lowest $p$ value at each genomic locus that reached nominal significance ($p$ < 5 x 10^-6^) indicated by the blue dashed line. The magenta pink highlighted SNPs indicate the locus across both GWAS that reached nominal significance in the merged GWAS. The dark orange highlighted SNPs indicate the locus that reached nominal significance in the GWAS meta-analysis.

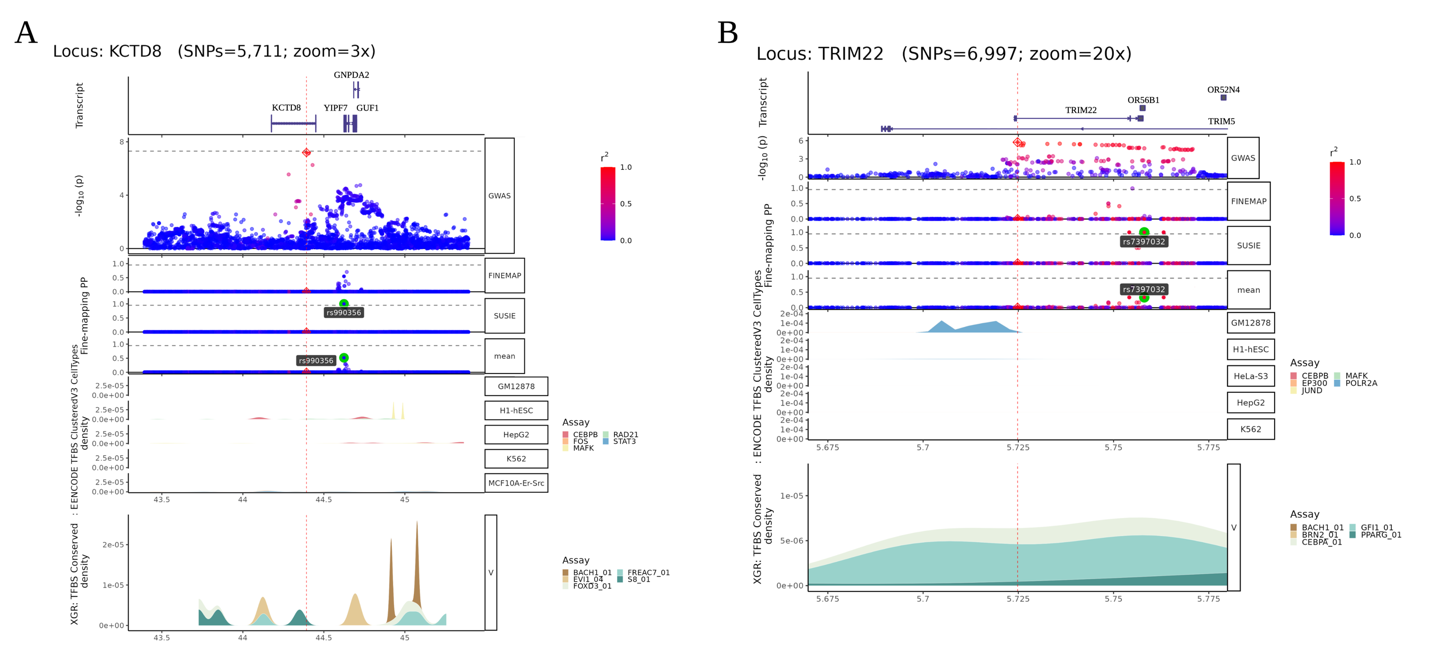

**Supplementary Figure 5 - Fine-mapping of the two lead loci**. (**A**) shows the chromosome 4 locus (*KCTD8*) and (**B**) shows the chromosome 11 (*TRIM22*) locus. Transcript plots are shown in the top row. The second row shows the GWAS results with the log_10_ $p$ value for each SNP (x-axis). The next three rows show the cross tools fine-mapping output (FINEMAP and SuSie) and the consensus results (mean) respectively. For the fine-mapping results the x-axis represents the per SNP posterior probability (PP). The final two rows show transcription factor binding site (TFBS) data for the region. Row six shows TFBS clustered by cell type from ENCODE and the final row the TFBS density conserved across species from XGR (human/mouse/rat).

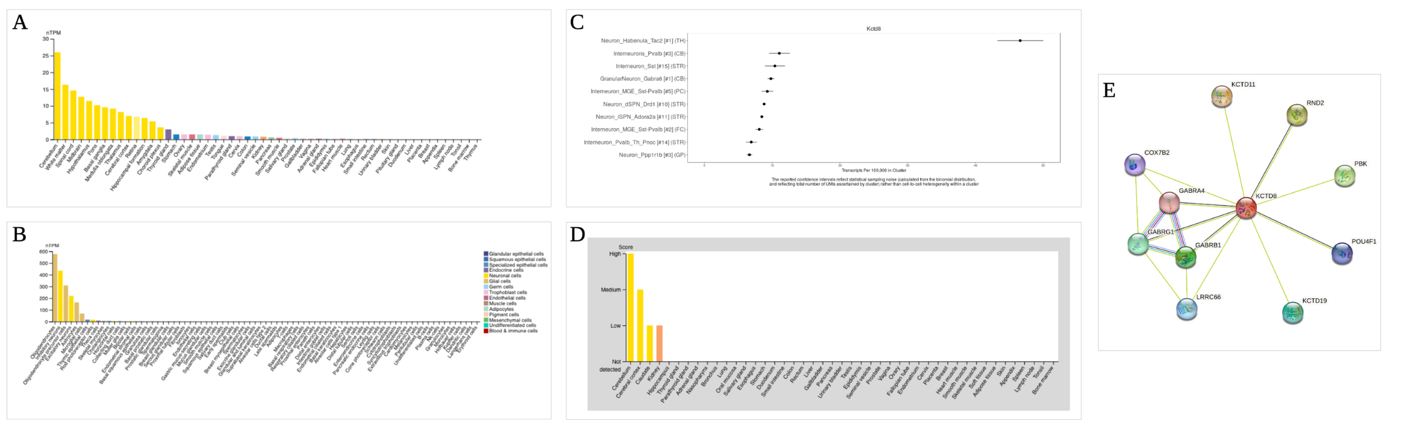

**Supplementary Figure 6 - *KCTD8* RNA and protein expression levels.** Comparison of (**A**) Bulk RNA expression across different tissue types (normalised transcripts per million (nTPM)) (**B**) single cell RNA (scRNA) expression (nTPM) across different cell types (**C**) Mouse scRNA expression (nTPM) across cell subtypes (**D**) Protein expression (score: high, medium, low or not detected on immunohistochemistry) by tissue type (**E**) Protein-protein interaction network of KCTD8 (predicted interactions: green lines = text mining, black = co-expression; known interactions: pink = experimentally determined, light blue = curated databases; purple = protein homology ). (**A-B**) Consensus data from Human Protein Atlas, (**C**) data from DropViz (**D**) IHC data from Human Protein Atlas (**E**) Protein interaction data from *string-db.org*

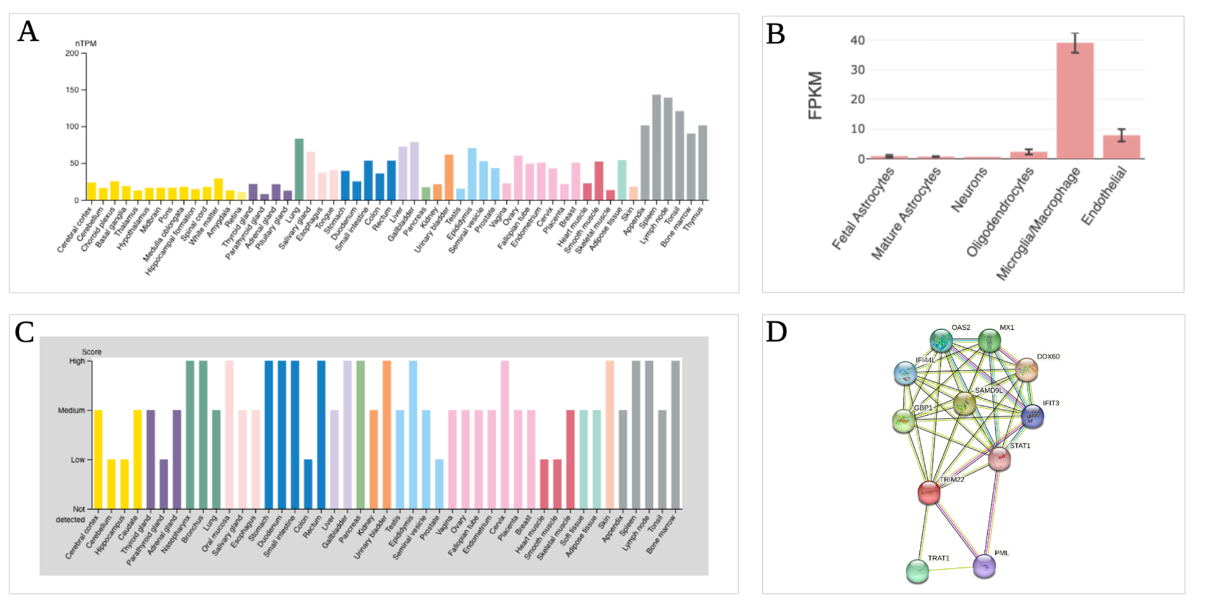

**Supplementary Figure 7 - *TRIM22* RNA and protein expression levels.** Comparison of **(A)** Bulk RNA expression across different tissue types (normalised transcripts per million (nTPM)) **(B)** Brain RNA expression (nTPM) across different cell types **(C)** Protein expression (score: high, medium, low or not detected on immunohistochemistry) by tissue type **(D)** Protein-protein interaction network of *TRIM22* (predicted interactions: green lines = text mining, black = co-expression; known interactions: pink = experimentally determined, light blue = curated databases; purple = protein homology ). **(A)** Consensus data from Human Protein Atlas, **(B)** data from BrainRNA-seq.org **(C)** IHC data from Human Protein Atlas **(D)** Protein interaction data from *string-db.org*

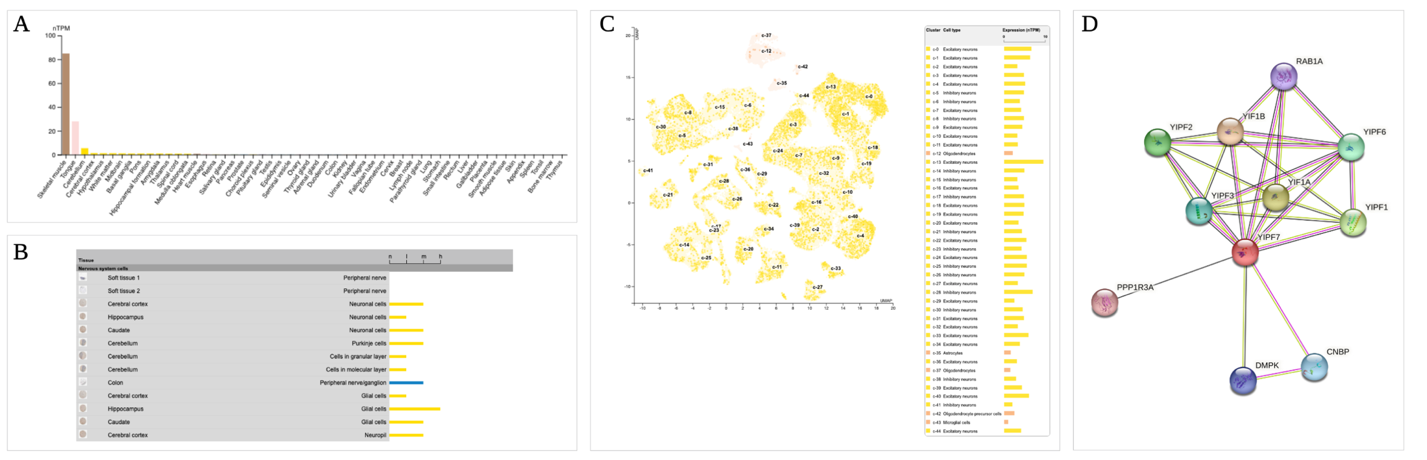

**Supplementary Figure 8 - *YIPF7* RNA and protein expression levels.** Comparison of **(A)** Bulk RNA expression across different tissue types (normalised transcripts per million (nTPM)) (**B**) Brain RNA expression (nTPM) across different cell types **(B)** Protein expression (score: high, medium, low or not detected on immunohistochemistry) by tissue type **(C)** Single cell RNA (scRNA) expression clusters for brain cell types **(D)** Protein-protein interaction network of YIPF7 (predicted interactions: green lines = text mining, black = co-expression; known interactions: pink = experimentally determined, light blue = curated databases; purple = protein homology ). **(A)** Consensus data from Human Protein Atlas, **(B)** IHC data from Human Protein Atlas **(C)** Consensus scRNA data from Human Protein Atlas **(D)** Protein interaction data from string-db.org.

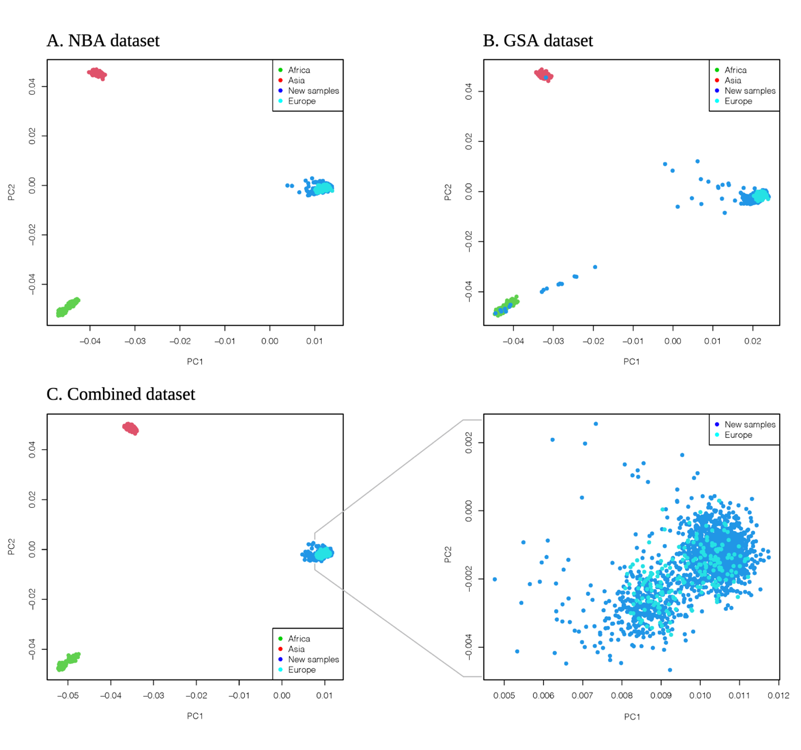

**Supp. Figure 9 - Genetic principal component plots.** First two principal components of the NBA (**A**) and GSA (**B**) and combined dataset (**C**) plotted against the HapMap3 Genome Reference Panel. These data include cases and controls. The NBA and GSA datasets are plotted before samples more than 6SD from the mean of the first 10 principal components were removed. For the combined dataset (**C**) all samples are within 6SD of the CEU reference population (outliers removed), which is expanded in the figure to the right.

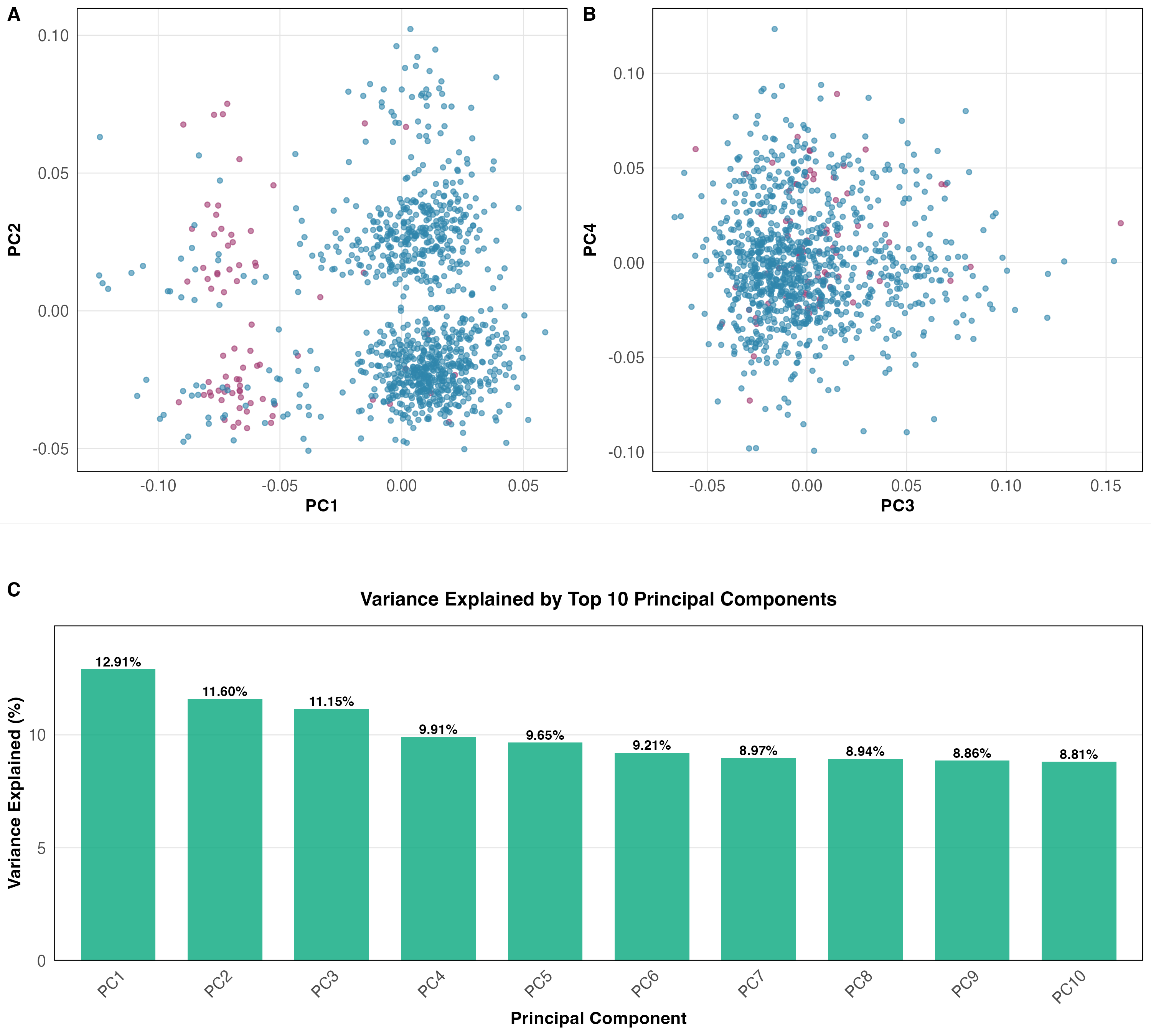

**Supplementary Figure 10:** Population stratification and genotyping array differences between GSA (in pink) and NBA (blue). A) Near complete separation of genotyping arrays in PC1 vs PC2. This is mostly limited to PC1 as B) PC3 vs PC4 does not replicate this separation. C) Variance explained by each PC.

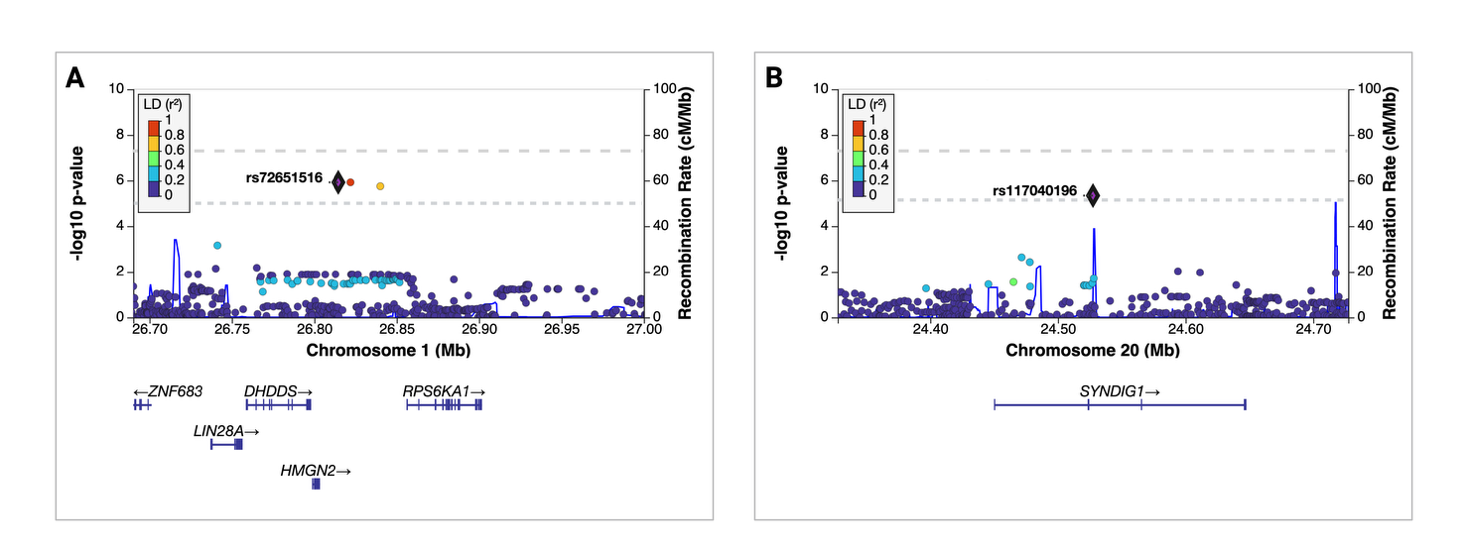

**Supplementary Figure 11: Regional association plots and recombination rates at suggestive genomic loci from meta-analysis.** Regional association plots at (A) 1:26814627 (rs72651516), (B) 20:24527458 (rs117040196). The index variants are indicated by a purple diamond and corresponding rsID. Linkage disequilibrium between the index variant and nearby variants, as measured by r2, is colour-coded (dark blue: 0 ≤ r2 < 0.2; light blue: 0.2 ≤ r2 < 0.4; green: 0.4 ≤ r2 < 0.6; orange: 0.6 ≤ r2 < 0.8; red: 0.8 ≤ r2 ≤ 1; blue: no r2 available). Genome-wide significance was set at $p$ < 5x10^-8^ and indicated by the top grey dashed line, while nominal significance (p < 5 x 10^-6^) is indicated by the lower grey dashed line All plots were generated in <http://locuszoom.org/>*.*

**(D) Supplementary Tables**

Supplementary Table 1 - Overview of total PiD samples included, broken down by contributing site.

| Contributing Site | Country | # samples |
| --- | --- | --- |
| UCL Institute of Neurology cohort (Europe / Australia) |  | 151 |
| Netherlands Brainbank, Amsterdam | Netherlands | 33 |
| UCL Queen Square Brain Bank, London | UK | 18 |
| Neurological Tissue Bank, Biobanc-Hospital Clínic-IDIBAPS, Barcelona | Spain | 14 |
| Manchester Brain Bank, Manchester | UK | 11 |
| Cambridge Brain Bank, Cambridge | UK | 11 |
| London Neurodegenerative Diseases Brain Bank, London | UK | 8 |
| South West Dementia Brain Bank, University of Bristol | UK | 8 |
| Sydney Brain Bank, Sydney | Australia | 8 |
| Neurobiobank München, Munich | Germany | 7 |
| Oxford Brain Bank, Oxford | UK | 7 |
| Newcastle Brain Tissue Resource, Newcastle-upon-Tyne | UK | 7 |
| Neuro-CEB Biobank France, Paris | France | 5 |
| Victorian Brain Bank, Florey Institute, Parkville, VIC | Australia | 5 |
| Douglas-Bell Canada Brain Bank, Montreal, QC | Canada | 3 |
| The Brain Bank at Karolinska Institutet, Stockholm | Sweden | 3 |
| Fondazione IRCCS Instituto Neurologico Carlo, Milan | Italy | 2 |
| DZNE Brain Bank, Tübingen | Germany | 1 |
| Mayo Clinic Jacksonville cohort (North America) |  | 170 |
| Mayo Clinic, Jacksonville, FL/Rochester, MN | USA | 55 |
| University of California, San Francisco, CA | USA | 21 |
| University of Pennsylvania, Philadelphia, PA | USA | 18 |
| Massachusetts General Hospital, Boston, MA | USA | 15 |
| Northwestern University, Chicago, IL | USA | 14 |
| Banner Sun Health Research, Sun City, AZ | USA | 9 |
| Sunnybrook Health Research, Toronto, ON | Canada | 9 |
| Columbia University, New York City, NY | USA | 9 |
| Emory University, Atlanta, GA | USA | 7 |
| University of British Columbia Hospital, Vancouver. BC | Canada | 5 |
| Houston Methodist Hospital, Houston. TX | USA | 4 |
| Krembil Research Institute, University of Toronto, Toronto, ON | Canada | 2 |
| UCLA - Sepulveda, Los Angeles, CA | USA | 1 |
| Dale E. Creighton Brain and BioBank, Western University, London, ON | Canada | 1 |
| Total PIC samples |  | 321 |

Supplementary Table 2: Summary of clinical diagnoses for Pick’s disease cases

| **Diagnosis** |  |  |  |  |  |  |
| --- | --- | --- | --- | --- | --- | --- |
| **Level 1** | **Level 2** | **Level 3** |  | **MCJ PiD cohort (GSA)** | **UCL PiD cohort (NBA)** | **Combined PiD cohort** |
| **FTD** | - | - |  | **126 (79.2%)** | **110 (81.5%)** | **236 (80.2%)** |
|  | bvFTD | - |  | 57 (35.8%) | 80 (59.3%) | 137 (46.5%) |
|  | PPA | - |  | 37 (23.3%) | 23 (17.0%) | 60 (20.4%) |
|  |  | nfvPPA |  | 13 (8.2%) | 8 (5.9%) | 21 (7.1%) |
|  |  | svPPA |  | 9 (5.7%) | 4 (3.0%) | 13 (4.4%) |
|  |  | not classified |  | 15 (9.4%) | 11 (8.1%) | 26 (8.8%) |
|  | not classified | - |  | 32 (20.1%) | 7 (5.2%) | 39 (13.3%) |
| **AD** | **-** | **-** |  | **19 (11.9%)** | **15 (11.1%)** | **34 (11.6%)** |
|  | Amnestic | - |  | 16 (10.1%) | 15 (11.1%) | 31 (10.5%) |
|  | lvPPA | - |  | 2 (1.3%) | 0 (0.0%) | 2 (0.7%) |
|  | PCA | - |  | 1 (0.6%) | 0 (0.0%) | 1 (0.3%) |
| **CBS** | - | - |  | **11 (6.9%)** | **3 (2.2%)** | **14 (4.8%)** |
| **Dementia not otherwise specified** | - | - |  | **2 (1.3%)** | **5 (3.7%)** | **7 (2.4%)** |
| **VaD** | - | - |  | **1 (0.6%)** | **1 (0.7%)** | **2 (0.7%)** |
| **No diagnosis** | - | - |  | **0 (0.0%)** | **1 (0.7%)** | **1 (0.3%)** |
| **Total** |  |  |  | **159 (100.0%)** | **135 (100.0%)** | **294 (100.0%)** |

**Abbreviations:** bvFTD = behavioural fronto-temporal dementia, CBS = Corticobasal syndrome, GSA = Illumina Global Screening Array, FTD = fronto-temporal dementia, lvPPA = logopenic variant primary progressive aphasia, MCJ = Mayo Clinic Jacksonville, NBA = Illumina NeuroBooster Array, nfvPPA = non-fluent variant primary progressive aphasia, PPA = primary progressive aphasia, PiD = Pick’s disease, svPPA = semantic variant primary progressive aphasia, UCL = University College London, VaD = Vascular dementia.

Supplementary table 3 - *MAPT* Haplotypes from GWAS

|  | H1/H1 | H1/H2 | H2/H2 |
| --- | --- | --- | --- |
| Cases | 47.6% (140) | 45.6% (134) | 6.8% (20) |
| Controls | 58.9% (621) | 36.1% (381) | 5.0% (53) |
| Total | 56.4% (761) | 38.2% (515) | 5.4% (73) |

Samples size (n) = 294 cases, 1055 controls within 6SD of CEU. MAPT haplotypes derived from rs8070723 H2 tagging SNP (minor allele G = H2). Chi square: $\chi^{2}$ = 6.04, df =2, $p$ = 0.003

Supplementary Table 4: Comparison of Top SNPs Across All Analytical Approaches

| Chr:Pos | rsID | Gene | Model | Beta (SE) | OR (95% CI) | P‑value |
| --- | --- | --- | --- | --- | --- | --- |
| **4:44392571** | **rs112161979** | ***KCTD8*** | Primary Combined (PC1,8,10) | 2.019 (0.373) | 7.53 (3.62‑15.65) | 6.37×10^⁻8^ |
|  |  |  | Combined (PC1-3) | 1.921 (0.368) | 6.83 (3.32‑14.06) | 1.82×10⁻⁷ |
|  |  |  | Interaction | 1.774 (0.363) | 5.89 (2.89‑12.00) | 1.01×10⁻⁶ |
|  |  |  | Metanalysis | 1.671 (0.374) | 5.31 (2.55‑11.06) | 8.20 x 10^-6^ |
|  |  |  | GSA‑Only | 2.352 (5.184) | 10.50 (0.0004‑268491) | 0.650 |
|  |  |  | NBA‑Only | 1.666 (0.370) | 5.29 (2.56‑10.93) | 6.81×10⁻⁶ |
| **11:5724803** | **rs66481907** | ***TRIM22*** | Primary Combined (PC1,8,10) | 0.740 (0.155) | 2.10 (1.54-2.84) | 1.83 x 10^-6^ |
|  |  |  | Combined (PC1-3) | 0.758 (0.155) | 2.13 (1.57‑2.89) | 1.09×10⁻⁶ |
|  |  |  | Interaction | 0.726 (0.160) | 2.07 (1.51‑2.83) | 5.36×10⁻⁶ |
|  |  |  | Metanalysis | 0.642 (0.168) | 1.90 (1.37‑2.64) | 1.38×10⁻⁴* |
|  |  |  | GSA‑Only | 0.887 (0.648) | 2.43 (0.68‑8.66) | 0.171 |
|  |  |  | NBA‑Only | 0.625 (0.171) | 1.87 (1.33‑2.61) | 2.66×10⁻⁴ |
| **15:27729149** | **rs112721576** | ***GABRG3*** | Primary Combined (PC1,8,10) | 1.315 (0.282) | 3.73 (2.14-6.48) | 3.10 x 10^-6^ |
|  |  |  | Combined (PC1-3) | 1.327 (0.282) | 3.77 (2.17‑6.55) | 2.46×10⁻⁶ |
|  |  |  | Interaction | 1.198 (0.277) | 3.31 (1.92‑5.70) | 1.53×10⁻⁵ |
|  |  |  | Metanalysis | 1.129 (0.288) | 3.09 (1.76-5.43) | 8.71×10⁻⁵ |
|  |  |  | GSA‑Only | 7.953 (2.845) | 2838 (10.8‑750000) | 0.005 |
|  |  |  | NBA‑Only | 1.058 (0.285) | 2.88 (1.65‑5.04) | 2.07×10⁻⁴ |
| **19:39029201** | **rs11881082** | ***RYR1*** | Primary Combined (PC1,8,10) | 1.085 (0.235) | 2.96 (1.87-4.69) | 4.00 x 10^-6^ |
|  |  |  | Combined (PC1-3) | 1.049 (0.232) | 2.86 (1.81‑4.50) | 6.17×10⁻⁶ |
|  |  |  | Interaction | 0.928 (0.230) | 2.46 (1.55-3.90) | 5.64×10⁻⁵ |
|  |  |  | Metanalysis | 0.900 (0.234) | 2.46 (1.55‑3.90) | 1.22×10⁻⁴ |
|  |  |  | GSA‑Only | ‑0.673 (1.241) | 0.51 (0.04‑5.81) | 0.588 |
|  |  |  | NBA‑Only | 0.959 (0.235) | 2.61 (1.65‑4.13) | 4.67×10⁻⁵ |
| **5:36376351** | **rs7720520** | ***RANBP3L*** | Primary Combined (PC1,8,10) | 0.562 (0.123) | 1.76 (1.38-2.23) | 4.50 x 10-6 |
|  |  |  | Combined (PC1-3) | 0.556 (0.122) | 1.74 (1.37‑2.22) | 5.61×10⁻⁶ |
|  |  |  | Interaction | 0.559 (0.128) | 1.75 (1.36‑2.25) | 1.28×10⁻⁵ |
|  |  |  | Metanalysis | 0.542 (0.136) | 1.72 (2.24-1.32) | 6.45×10⁻⁵ |
|  |  |  | GSA‑Only | 1.446 (0.650) | 4.25 (1.19‑15.19) | 0.026 |
|  |  |  | NBA‑Only | 0.501 (0.137) | 1.65 (1.26‑2.16) | 2.52×10⁻⁴ |

Combined model (PC1,2,8) – Primary Combined Model; adjusted for age, array, sex, and 3 principal components (PC1, PC8, PC10). All complimentary models adjusted for age, sex, and top 3 principal components (PC1, PC2, PC3). Combined model (PC1-3) also adjusted for array. Each model was used to study the association between 6,316,457 variants and risk of PiD (294 cases, 1055 controls). Abbreviations: Beta=Natural log of (OR), OR = odds ratio, SE = standard error, CI = confidence interval. *the lead SNP rs7936441 (OR 1.94, 95% CI 1.40 – 2.69, p = 6.47 x 10^-5^) at Chromosome 11 (TRIM22) locus in the meta-analysis is in complete linkage disequilibrium with rs66481907.

Supplementary Table 5 - Fine-mapping results of *KCTD8* and *TRIM22* loci

| SNP | CHR | N | T_stat | FINEMAP.CS | FINEMAP.PP | SUSIE.CS | SUSIE.PP | Support | Consensus | Mean.PP |
| --- | --- | --- | --- | --- | --- | --- | --- | --- | --- | --- |
| rs990356 | 4 | 2668 | 3.80 | 1 | 0 | 2 | 1 | 1 | FALSE | 0.33 |
| rs11038885 | 11 | 2692 | 4.32 | 1 | 0 | 3 | 1 | 1 | FALSE | 0.25 |
| rs7397032 | 11 | 2688 | 4.29 | 1 | 0 | 2 | 1 | 1 | FALSE | 0.25 |
| rs76253329 | 11 | 2678 | 2.26 | 1 | 0.96697 | 0 | 0 | 1 | FALSE | 0.15 |

Abbreviation = N, Sample size for fine-mapping; t_stat = test statistic; CS = Credible Set; PP = Posterior Probability. Mean.PP = the mean posterior probability from the two fine-mapping posterior probabilities

Supplementary Table 6 - Variance Explained by Top 10 Principal Components and their association with GSA.

| PC | PC1 | PC2 | PC3 | PC4 | PC5 | PC6 | PC7 | PC8 | PC9 | PC10 |
| --- | --- | --- | --- | --- | --- | --- | --- | --- | --- | --- |
| Variance Explained (%) | 2.84 | 1.92 | 1.47 | 1.21 | 1.08 | 0.95 | 0.87 | 0.79 | 0.72 | 0.68 |
| Association with Array  ($p$ value) | 8.3 × 10⁻²⁸ | 1.2 × 10⁻⁸ | 0.003 | 0.142 | 2.1 × 10⁻⁵ | 0.089 | 1.3 × 10⁻⁴ | 0.251 | 0.417 | 0.001 |

Supplementary Table 7 - Top independent SNPs at suggestive loci from PiD GWAS Metanalysis

| Chr | BP | SNP | Nearest gene | Minor allele | OR (95% CI) | $\boldsymbol{p}$ value |
| --- | --- | --- | --- | --- | --- | --- |
| 1 | 26,814,627 | rs72651516 | HMGN2 | A | 2.94 (1.90 – 4.55) | 1.20 x 10^-6^ |
| 20 | 24,527,458 | rs117040196 | SYNDIG1 | G | 5.10 (2.54-10.2) | 4.57 x 10^-6^ |

Logistic regression additive model adjusted for gender, age, array and top three within-cohort PCs conducted separately on the NBA and GSA cohorts. Each model was used to study the association between 6,316,457 variants and risk of PiD (294 cases, 1055 controls). Resulting summary statistics then combined in a fixed-effects meta-analysis using *metal*. Abbreviations: BP = base-pair coordinate, Chr = chromosome, CI = confidence interval, OR = odds ratio, SNP = single nucleotide polymorphism.

#### **(E) Pick’s Disease International Consortium (PIC)**

**Members**

Matthew C Baker^1^, Alexandra I Soto-Beasley^1^, Professor. Tamas Revesz^2,3^, Professor. Thomas T Warner^2,4^, Professor. Zane Jaunmuktane^2,4^, Professor. Bradley F Boeve^5^, Elizabeth A Christopher^1^, Michael DeTure^1^, Professor. Ranjan Duara^6^, Professor. Neill R Graff-Radford^7^, Professor. Keith A Josephs^5^, Shunsuke Koga^1^, Professor. Melissa E Murray^1,8^, Professor. Kelly E Lyons^9^, Professor. Rajesh Pahwa^9^, Professor. Ronald C Petersen^5^, Professor. Jennifer L Whitwell^10^, Professor. Lea T Grinberg^11^, Professor. Bruce Miller^11^, Athena Schlereth^11^, Professor. William W Seeley^11^, Professor. Salvatore Spina^11^, Professor. David J Irwin^12^, Professor. Edward B Lee^13^, EunRan Suh^13^, Professor. Vivianna M Van Deerlin^13^, Professor. David A Wolk^12^, Theresa R Connors^14^, Professor. Matthew P Frosch^14^, Derek H Oakley^14^, Iban Aldecoa^15^, Mircea Balasa^16,15^, Professor. Ellen Gelpi^17^, Sergi Borrego-Écija^16,15^, Rosa Maria de Eugenio Huélamo^18^, Jordi Gascon-Bayarri^19^, Professor. Raquel Sánchez-Valle^16,15^, Pilar Sanz-Cartagena^20^, Gerard Piñol-Ripoll^21^, Laura Molina-Porcel^15,16,22^, Margaret E Flanagan^23,24^, Tamar Gefen^23,25^, Emily J Rogalski^23,25,26,27^, Professor. Sandra Weintraub^23,25^, Professor. Julie A Schneider^28^, Xiongwei Zhu^29^, Javier Redding-Ochoa^30^, Koping Chang^30^, Professor. Juan C Troncoso^30^, Stefan Prokop^31^, Professor. Kathy L Newell^32^, Professor. Bernardino Ghetti^32^, Max Jacobsen^33^, Matthew Jones^34,35^, Anna Richardson^34,35^, Andrew C Robinson^36,37^, Professor. Federico Roncaroli^36,38^, Professor. Julie Snowden^34,35^, Kieren Allinson^39^, Annelies Quaegebeur^40,39^, Professor. Thomas G Beach^41^, Geidy E Serrano^41^, Xena E Flowers^42^, Professor. James E Goldman^43^, Allison C Heaps^42^, Sandra P Leskinen^42^, Andrew F Teich^44,42^, Professor. Sandra E Black^45^, Julia L Keith^46^, Mario Masellis^45^, Istvan Bodi^47,48^, Andrew King^47,48^, Professor. Safa-Al Sarraj^47,48^, Claire Troakes^48^, Professor. Glenda M Halliday^49^, Professor. John R Hodges^49^, Professor. Jillian J Kril^50^, Professor. John B Kwok^49^, Professor. Olivier Piguet^51^, Marla Gearing^52^, Sigrun Roeber^53^, Thomas Arzberger^54^, Professor. Jochen Herms^55,56,57^, Professor. Johannes Attems^58^, Christopher M Morris^58^, Professor. Alan J Thomas^58^, Bret M. Evers^59^, Professor. Charles L White, 3rd^59^, Kevin F Bieniek^60,61^, Professor. Naguib Mechawar^62^, Anne A Sieben^63,64,65,66^, Professor. Patrick P Cras^63,64,67^, Bart B De Vil^63,64,67^, Professor. Peter P De Deyn^68^, Professor. Isabelle Le Ber^69,70^, Professor. Danielle Seilhean^71^, Susana Boluda^71^, Sabrina Turbant-Leclere^72^, Professor. Ian R MacKenzie^73^, Professor. Catriona McLean^74,75^, Matthew D Cykowski^76^, John F Ervin^77^, Shih-Hsiu J Wang^78^, Professor. Caroline Graff^79,80^, Inger Nennesmo^81,82^, Rashed M Nagra^83^, James Riehl^84^, Professor. Gabor G Kovacs^85,86^, Giorgio Giaccone^87^, Benedetta Nacmias^88,89^, Professor. Manuela Neumann^90,91^, Professor. Lee-Cyn Ang^92,93^, Elizabeth C Finger^94,95^, Cornelis Blauwendraat^96^, Mike A Nalls ^97,98,99^, Professor. Andrew B Singleton^96^, Dan Vitale^97,98,99^, Cristina Cunha^100^, Agostinho Carvalho^100,101^, Professor. Zbigniew K Wszolek^7^, C. Dirk Keene^102^, Caitlin S Latimer^102^, Aimee Schantz^102^, Thomas D Bird^103,104,105^, Vahram Haroutunian^106^, Dushyant Purohit^106^, Jamie M Walker^106^, Jeffrey Metcalf^107^, Robert Rissman^108^, Professor. Richard J Perrin^109^, Erin E Franklin^109^, Ann M Chaffee^109^

**Affiliations**

^1^Department of Neuroscience, Mayo Clinic, Jacksonville, FL 32224, USA, ^2^Queen Square Brain Bank for Neurological Disorders, University College London, Queen Square Institute of Neurology London, UK, ^3^Department of Neurodegenerative Disease, University College London, Queen Square Institute of Neurology, London, UK, ^4^Department of Clinical and Movement Neurosciences, University College London, Queen Square Institute of Neurology, London, UK, ^5^Department of Neurology, Mayo Clinic, Rochester, MN 55905, USA, ^6^Wien Center for Alzheimer’s Disease and Memory Disorders, Mount Sinai Medical Center Miami Beach, FL, ^7^Department of Neurology, Mayo Clinic, Jacksonville, FL 32224, USA, ^8^Department of Laboratory Medicine and Pathology, Mayo Clinic, Jacksonville, FL 32224, USA, ^9^University of Kansas Medical Center, Parkinson’s Disease & Movement Disorder Division, Kansas City, KS. 66160, ^10^Department of Radiology, Mayo Clinic, Rochester, MN 55905, USA, ^11^Department of Neurology, Memory and Aging Center, University of California San Francisco, San Francisco, CA, USA, ^12^Department of Neurology, Perelman School of Medicine at the University of Pennsylvania, Philadelphia, PA 19104, USA, ^13^Department of Pathology and Laboratory Medicine, Perelman School of Medicine at the University of Pennsylvania, Philadelphia, PA 19104, USA, ^14^Neuropathology Service, C.S. Kubik Laboratory for Neuropathology, Massachusetts General Hospital/Harvard Medical School, Boston, MA, USA, ^15^Neurological Tissue Bank, Biobanc–Hospital Clínic, Clinical Research Foundation–August Pi i Sunyer Biomedical Research Institute (FRCB-IDIBAPS), Barcelona, Spain, ^16^Alzheimer’s Disease and Other Cognitive Disorders Unit, Neurology Service, Hospital Clínic, FRCB-IDIBAPS, and Institute of Neurosciences, University of Barcelona, Barcelona, Spain, ^17^Division of Neuropathology and Neurochemistry, Department of Neurology, Medical University of Vienna, Vienna, Austria, ^18^Hospital de Palamós, Carrer Hospital, 36, 17230 Palamós, Girona, Spain, ^19^Servei de Neurologia, Hospital Universitari de Bellvitge. Institut d'Investigació Biomèdica de Bellvitge (Idibell). L’Hospitalet de Llobregat, Spain, ^20^Hospital de Mataro, Carr. de Cirera, 230, 08304 Mataró, Barcelona, Spain, ^21^Unitat Trastorns Cognitius (Cognitive Disorders Unit), Clinical Neuroscience Research, IRBLleida, Santa Maria University Hospital, Lleida, Spain, ^22^Center of Biomedical Research Network for Neurodegenerative Diseases (CIBERNED), Madrid, Spain, ^23^Mesulam Center for Cognitive Neurology & Alzheimer’s Disease, Northwestern University Feinberg School of Medicine, Chicago, IL, USA, ^24^Department of Pathology, Northwestern University Feinberg School of Medicine, Chicago, IL, USA, ^25^Department of Psychiatry and Behavioral Sciences, Northwestern University Feinberg School of Medicine, Chicago, IL, USA, ^26^Healthy Aging & Alzheimer's Research Care Center, Biological Sciences Division, University of Chicago, IL, ^27^Department of Neurology, Biological Sciences Division, University of Chicago, IL, ^28^Rush Alzheimer's Disease Center, Rush University Medical Center, Chicago, IL, USA, ^29^Department of Pathology, Case Western Reserve University, 2103 Cornell Road, Cleveland, OH, 44106, USA, ^30^Johns Hopkins School of Medicine, Baltimore, MD, USA, ^31^Fixel Institute for Neurological Diseases, University of Florida, Gainesville, FL, USA, ^32^Department of Pathology and Laboratory Medicine, Indiana University School of Medicine, Indianapolis, Indiana 46202, USA, ^33^Department of Pathology and Laboratory Medicine, Indiana University School of Medicine, Indianapolis, IN, USA, ^34^Cerebral Function Unit, Manchester Centre for Clinical Neurosciences, Salford Royal NHS Foundation Trust, UK, ^35^Division of Neuroscience, School of Biological Sciences, University of Manchester, UK, ^36^Division of Neuroscience, Faculty of Biology, Medicine and Health, School of Biological Sciences, The University of Manchester, Salford Royal Hospital, Salford, M6 8HD, UK, ^37^Geoffrey Jefferson Brain Research Centre, Manchester Academic Health Science Centre (MAHSC), Manchester, UK., ^38^Geoffrey Jefferson Brain Research Centre, Manchester Academic Health Science Centre (MAHSC), Manchester, UK, ^39^Cambridge University Hospitals NHS Trust, Cambridge CB2 00Q, UK, 39 Department of Clinical Neurosciences, University of Cambridge, UK, ^40^Department of Clinical Neurosciences, University of Cambridge, UK, ^41^Civin Laboratory of Neuropathology, Banner Sun Health Research Institute, Sun City, AZ 85351, USA, ^42^Taub Institute for Research on Alzheimer’s Disease and the Aging Brain, Columbia University, New York, NY, USA, ^43^Department of Pathology and Cell Biology, Columbia University Irving Medical Center, Vagelos College of Physicians and Surgeons, Columbia University, New York, NY 10032, USA, ^44^Department of Pathology and Cell Biology, Columbia University, New York, NY, USA, ^45^Department of Medicine, Division of Neurology, Sunnybrook Health Sciences Centre and University of Toronto, Hurvitz Brain Sciences Research Program, Sunnybrook Research Institute, ^46^Laboratory Medicine and Molecular Diagnostics, Sunnybrook Health Sciences Centre, and Laboratory Medicine and Pathobiology, University of Toronto, ^47^Clinical Neuropathology Department, King's College Hospital NHS Foundation Trust, London, UK, ^48^London Neurodegenerative Diseases Brain Bank, Department of Basic and Clinical Neuroscience, Institute of Psychiatry, Psychology and Neuroscience, King's College London, London, UK, ^49^University of Sydney Brain and Mind Centre and Faculty of Medicine and Health School of Medical Sciences, ^50^University of Sydney Faculty of Medicine and Health School of Medical Sciences, ^51^University of Sydney Brain and Mind Centre and Faculty of Science School of Psychology, ^52^Dept. of Pathology and Laboratory Medicine, Dept. of Neurology, and Goizueta Alzheimer's Disease Center Brain Bank; Emory University School of Medicine, Atlanta, GA USA, ^53^Center for Neuropathology and Prion Research, Ludwig-Maximilians-University Munich, Germany, ^54^Department of Psychiatry and Psychotherapy, University Hospital, Ludwig-Maximilians-University Munich, Germany, ^55^German Center for Neurodegenerative Diseases (DZNE), Munich, Germany, ^56^Center for Neuropathology and Prion Research, Ludwig-Maximilians-Universität, Munich, Germany, ^57^Munich Cluster for Systems Neurology (Synergy), Munich, Germany, ^58^Newcastle Brain Tissue Resource, NIHR Biomedical Research Centre (BRC) Newcastle, Translational and Clinical Research Institute, Edwardson Building, Newcastle University, Newcastle upon Tyne, NE4 5PL, UK, ^59^University of Texas Southwestern Medical Center, Dallas, TX 75390, ^60^Glenn Biggs Institute for Alzheimer's and Neurodegenerative Diseases, San Antonio, TX, USA, ^61^University of Texas Health San Antonio, San Antonio, TX, USA, ^62^Douglas Hospital Research Centre, McGill University, Montreal, QC, Canada, ^63^Laboratory of Neurology, Translational Neurosciences, Faculty of Medicine and Health Sciences, University of Antwerp, Antwerp, Belgium, ^64^IBB-NeuroBiobank BB190113, Born Bunge Institute, Antwerp, Belgium, ^65^Department of Pathology, Antwerp University Hospital, Antwerp, Belgium, ^66^Department of Neurology, Ghent University Hospital , Ghent University, Belgium, ^67^Department of Neurology, Antwerp University Hospital - UZA, Antwerp, Belgium, ^68^Laboratory of Neurochemistry and Behavior, Experimental Neurobiology Unit, University of Antwerp, Universiteitsplein 1, 2610 Antwerpen, Belgium, ^69^Inserm U1127, CNRS UMR 7225, Sorbonne Université, Paris Brain Institute (ICM), Hôpital Pitié-Salpêtrière, Paris, France, ^70^Centre de référence des démences rares ou précoces, Hôpital Pitié-Salpêtrière, Paris, France, ^71^Laboratoire de Neuropathologie Escourolle, Hôpital de la Salpêtrière, AP-HP, & Alzheimer Prion Team, ICM, 47 Bd de l'Hôpital, 75651 CEDEX 13 Paris, France, ^72^Inserm U1127, CNRS UMR 7225, Sorbonne Université, Paris Brain Institute (ICM) Hôpital Pitié-Salpêtrière, Paris, France, ^73^Department of Pathology and Laboratory Medicine, University of British Columbia, Vancouver, BC Canada V6T 2B5, ^74^Department of Anatomical Pathology Alfred Heath, Melbourne, Victoria, 3004, Australia, ^75^Victorian Brain Bank, The Florey Institute of Neuroscience of Mental Health, Parkville, Victoria, 3052, Australia, ^76^Department of Pathology and Genomic Medicine, Houston Methodist Research Institute and Weill Cornell Medicine, Houston, TX, ^77^Department of Neurology, Duke University Medical Center, Durham, USA, ^78^Department of Pathology, Duke University Medical Center, Durham, USA, ^79^Division for Neurogeriatrics, Centre for Alzheimer Research, Department of Neurobiology, Care Sciences and Society, Karolinska Institutet, Stockholm, Sweden, ^80^Unit for Hereditary Dementias, Karolinska University Hospital Solna, Stockholm, Sweden, ^81^Dept of Oncology-Pathology, Karolinska Institutet, Stockholm, ^82^Dept of Pathology and Cancer Diagnostics, Karolinska University Hospital Solna, Stockholm, Sweden, ^83^Human Brain and Spinal Fluid Resource Center, Brentwood Biomedical Research Institute, Los Angeles, CA, United States, ^84^UCLA - Sepulveda, Los Angeles, CA, ^85^Tanz Centre for Research in Neurodegenerative Disease (CRND) and Department of Laboratory Medicine and Pathobiology, University of Toronto, Toronto, ON, Canada, ^86^Laboratory Medicine Program and Krembil Brain Institute, University Health Network, Toronto, ON, Canada, ^87^Fondazione IRCCS Istituto Neurologico Carlo Besta, Milan, Italy, ^88^Department of Neuroscience, Psychology, Drug Research and Child Health University of Florence, Florence, Italy, ^89^IRCCS Fondazione Don Carlo Gnocchi, Florence, Italy, ^90^Molecular Neuropathology of Neurodegenerative Diseases,German Center for Neurodegenerative Diseases (DZNE), Tübingen, Germany, ^91^Department of Neuropathology, University Hospital of Tübingen, Tübingen, Germany, ^92^Department of Pathology and Laboratory Medicine, London Health Sciences Centre, London, ON, Canada, ^93^Schulich School of Medicine and Dentistry, Western University, London. ON, Canada, ^94^Department of Clinical Neurological Sciences, Western University, London, ON, Canada, ^95^Schulich School of Medicine and Dentistry, Western University, London, ON, Canada, ^96^Global Parkinson's Genetics Program, Chevy Chase, MD, USA, ^97^Center for Alzheimer’s and Related Dementias, Bethesda, MD, USA 20892, ^98^DataTecnica, Washington, DC, USA 20037, ^99^Laboratory of Neurogenetics, NIA, NIH, Bethesda, MD, USA 20892, ^100^Life and Health Sciences Research Institute (ICVS), School of Medicine, University of Minho, Braga, Portugal, ^101^ICVS/3B’s - PT Government Associate Laboratory, Braga/Guimarães, Portugal, ^102^Department of Laboratory Medicine and Pathology, University of Washington School of Medicine, ^103^Geriatrics Research Education and Clinical Center, Veterans Affairs Puget Sound Health Care System, Seattle, WA, USA, ^104^Department of Neurology, University of Washington, Seattle, Washington, USA, ^105^Division of Medical Genetics, Department of Medicine, University of Washington, Seattle, WA, USA, ^106^Icahn School of Medicine at Mount Sinai, ^107^University of California, San Diego, School of Medicine, ^108^Alzheimer's Therapeutic Research Institute Keck School of Medicine of the University of Southern California San Diego California USA., ^109^Department of Pathology and Immunology, Department of Neurology, Washington University School of Medicine, Saint Louis, Missouri, 63110.

**Acknowledgements**

*Mayo Clinic:* SK receives funding from CurePSP and the Rainwater Charitable Foundation, the State of Florida Ed and Ethel Moore Alzheimer's Disease Research Program (22A05), and Mayo Clinic Alzheimer's Disease Research Center (ADRC). MEM receives funding from the State of Florida (20A22), LEADS Neuropathology Core (U01AG057195), and the Chan Zuckerberg Initiative Collaborative Pairs Grant, which are paid directly to the institute. KAJ is supported by National Institutes of Health (NIH) grants (R01 DC014942, R01, R01-AG37491, R01-NS89757, RF1-NS112153 and RF1-NS120992). BFB is supported by National Institutes of Health (NIH) grants (P30 AG62677, U01 NS100620, R34 AG056639); the Robert H. and Clarice Smith and Abigail Van Buren Alzheimer s Disease Research Program of the Mayo Foundation; the Lewy Body Dementia Association; the Mayo Clinic Dorothy and Harry T. Mangurian Jr. Lewy Body Dementia Program; the Little Family Foundation; the Turner Foundation. ZKW is partially supported by the NIH/NIA and NIH/NINDS (1U19AG063911), the Haworth Family Professorship in Neurodegenerative Diseases fund, and the gifts from The Albertson Parkinson's Research Foundation and the Margaret M. and John Wilchek Family. ZKW serves as Mayo Clinic PI on Amylyx AMX0035-009 project, as Co-PI of the Mayo Clinic APDA Center for Advanced Research, and as an external advisory board member for the Sanannah Biotherapeutics, Inc. KAJ and JLW receive research support from the NIH (R01-DC12519, R01-NS89757, R01-AG50603, R01-DC14942, R01-AG37491, RF1-NS112153, and RF1-NS120992). Samples included in this study were clinical controls from Mayo Clinic Rochester and Mayo Clinic Jacksonville as part of the Alzheimer’s Disease Research Center (P30 AG062677), and the Mayo Clinic Study of Aging (U01 AG006786) or tissue donations to the Mayo Clinic Brain Bank in Jacksonville which is supported by CurePSP and Mayo Clinic funding.

*UCSF*: Human tissue samples were provided by the Neurodegenerative Disease Brain Bank at the University of California, San Francisco, which receives funding support from NIH grants P01AG019724 and P50AG023501, the Consortium for Frontotemporal Dementia Research, and the Rainwater Charitable Foundation. LTG and SS receive funding from NIH grants K24053435 and K08AG052648, respectively.

*UPenn*: ES, MG, VMVD, DJI, DAW, and EBL all receive funding through NIH - ES: P01-AG017586, P01-AG066597, P30-AG010124, P30-AG072979; MG: P01-AG017586, P01-AG066597, P30-AG010124, P30-AG072979; VMVD: P01-AG017586, P01-AG066597, P30-AG010124, P30-AG072979; DJI: R01-NS109260, P30-AG010124, P01-AG066597; DAW: P30-AG010124, P30-AG072979; and EBL: P01-AG017586, P01-AG066597, P30-AG010124, P30-AG072979.

*Northwestern*: ER, TG, SW, EHB, MEF receive support from NIA under award numbers R01 AG062566, R01 AG077444, P30 AG13854, P30 AG072977; the National Institute of Deafness and Other Communication Disorders (NIDCD) under award number R01 DC008552; and the National Institute of Neurological Disorders and Stroke (NINDS) under award number R01 NS075075. MEF also receives support from NIA grant K08 AG065463.

*Banner*: We are grateful to the Banner Sun Health Research Institute Brain and Body Donation Program of Sun City, Arizona for the provision of human biological materials. The Brain and Body Donation Program has been supported by the NINDS (U24 NS072026 National Brain and Tissue Resource for Parkinson’s Disease and Related Disorders), the NIA (P30 AG019610 and P30AG072980, Arizona Alzheimer’s Disease Core Center, the Arizona Department of Health Services (contract 211002, Arizona Alzheimer’s Research Center), the Arizona Biomedical Research Commission (contracts 4001, 0011, 05-901 and 1001 to the Arizona Parkinson's Disease Consortium), and the Michael J. Fox Foundation for Parkinson’s Research.

*UF:* The UF HBTB is supported by grants from the National Institute on Aging (P30AG066506, P50AG047266) and funds from the McKnight Brain institute (MBI).

*UT Southwestern:* CLW is supported by NIH Grant P30 AG012300; McCune Foundation; Winspear Family Center for Research on the Neuropathology of Alzheimer Disease

*UW:* TDB is supported by NIH grant K08 AG065426

*Icahn:* VH is supported by NIH grants (NIH-75N95019C00049, P30AG066514), DP by NIH (P30AG066514) and JMW by NIH P30 AG066509 (UW ADRC) and the Nancy and Buster Alvord Endowment.

*Emory*: MG is supported by NIH grant P30 AG066511. MDC receives funding from NIH grant RF1 NS118584.

*Columbia*: We thank the Columbia University Alzheimer's Disease Research Center (ADRC), funded by NIH grant P30AG066462, to S.A. Small (P.I.), and A. Teich from New York Brain Bank for providing biological samples and associated information. The ADRC is supported by the National Institutes of Health, through grant number P30AG066462. ACH also receives NIH support through P30AG066462.

*Duke:* The Bryan Brain Bank and Biorepository of the Duke-UNC ADRC and SJW are supported by the NIA grant P30AG072958

*MGH:* Brain samples were provided by Neuropathology Core of the Massachusetts Alzheimer Disease Research Center, which receives funding support from NIH grant P30 AG062421, which also supported TRC, PMD, MPF and DHO. DHO was also received support from the Dr. and Mrs. E. P. Richardson, Jr, Fellowship in Neuropathology.

*Indiana:* BG is supported by the US National Institutes of Health (grant P30-AG010133).

*NIH:* This research was supported by the Intramural Research Program, National Institute on Aging, National Institutes of Health, Department of Health and Human Services, project ZO1 AG000949. This work utilized the computational resources of the NIH STRIDES Initiative (https://cloud.nih.gov) through the Other Transaction agreement - Azure: OT2OD032100, Google Cloud Platform: OT2OD027060, Amazon Web Services: OT2OD027852. This work utilized the computational resources of the NIH HPC Biowulf cluster (https://hpc.nih.gov). Disclosure H.L.L. and M.A.N's participation in this project was part of a competitive contract awarded to DataTecnica by the National Institutes of Health (NIH) to support open science research. Some authors’ participation in this project was part of a competitive contract awarded to DataTecnica by the National Institutes of Health to support open science research. M.A.N. also owns stock in Character Bio Inc. and Neuron23 Inc.

*Washington University in Saint Louis:* We thank the participants and personnel of the Knight Alzheimer's Disease Research Center, the staff and patients of the Washington University Memory Diagnostic Center, and the staff of Washington University’s Translational Human Neurodegenerative Disease Research (THuNDR) Laboratory for providing postmortem human brain tissue samples for this study. The Knight Alzheimer Disease Research Center is supported by grants P01 AG003991 (Healthy Aging and Senile Dementia), P30 AG066444 (Alzheimer’s Disease Research Center) and P01 AG026276 (Adult Children Study). R.J.P., E.E.F. and A.M.C. are supported by NIH grants R01AG068319, R01 AG053267, P01 AG003991, P30 AG066444, U19 AG024904, U19 AG032438, U19 AG07879, U19 AG069701, RF1NS139970, R01 AG074909, R01 AG058676, R01 AG054567, R01 AG052550, R01NS097799, R01NS092865, and R01 NS075321, by the American Parkinson Disease Association, and by Target ALS. R.J.P. and E.E.F. are additionally supported by R01 NS134586. R.J.P. is additionally supported by R01 AG070883 and U19 NS110456. R.J.P.'s THuNDR Laboratory receives cost recovery funding from Biogen for tissue procurement and processing services related to ALS clinical trials. Neither R.J.P., E.E.F., A.M.C., nor their families own stock or have equity interest (outside of mutual funds or other externally directed accounts) in any pharmaceutical or biotechnology company.

*Western University*: We also acknowledge the DEC Brain & Biobank.

*Krembil*: GGK receives funding from The Rossy Foundation and Edmond J. Safra Philanthropic Foundation.

*Dale E. Creighton Brain and BioBank:* with thanks for contribution of samples to the study.

*McGill*: The Douglas-Bell Canada Brain Bank is funded by Healthy Brains for Healthy Lives (CFREF), the Réseau Québécois sur le suicide, le troubles de l’humeur et les troubles associés (FRQ-S), and by Brain Canada. NM is funded by a CIHR project grant.

*KCL:* The London Neurodegenerative Diseases Brain Bank, KCL, receives funding from the MRC and as part of the Brains for Dementia Research project (jointly funded by the Alzheimer’s Society and Alzheimer’s Research UK).

*Cambridge*: Cambridge Brain Bank is supported by the NIHR Cambridge Biomedical Research Centre. JBR receives support from Wellcome Trust (220258) and NIHR Cambridge Biomedical Research Centre (BRC-1215-20014). The views expressed are those of the authors and not necessarily those of the NIHR or the Department of Health and Social Care; PSP Association and Evelyn Trust; Medical Research Council (SUAG051 R101400).

*Bristol*: We would like to thank the South West Dementia Brain Bank (SWDBB), their donors and donor’s families for donating brain tissue for this study. Tissue for this study was provided with support from the BDR programme, jointly funded by Alzheimer's Research UK and Alzheimer's Society, and BRACE.

*Oxford*: We acknowledge the Oxford Brain Bank, supported by the Medical Research Council (MRC), Brains for Dementia Research (BDR) (Alzheimer Society and Alzheimer Research UK), Autistica UK, and the NIHR Oxford Biomedical Research Centre.

*Newcastle*: Tissue for this study was provided by the Newcastle Brain Tissue Resource which is funded in part by a grant from the UK Medical Research Council (G0400074), by NIHR Newcastle Biomedical Research Centre awarded to the Newcastle upon Tyne NHS Foundation Trust and Newcastle University, and as part of the Brains for Dementia Research Programme jointly funded by Alzheimer’s Research UK and Alzheimer’s Society.

*Manchester:* Tissue samples were supplied by The Manchester Brain Bank, which is part of the Brains for Dementia Research programme, jointly funded by Alzheimer’s Research UK and Alzheimer’s Society.

*Barcelona*: We are indebted to the HCB-IDIBAPS Biobank, integrated in the Spanish National Biobanks Network, for the biological human samples and data procurement. Gerard Piñol-Ripoll acknowledges the support from the Department of Health (PERIS 2019 SLT008/18/00050). Sergi Borrego-Écija is funded by the Joan Rodés - Josep Baselga grant from the FBBVA.

*Amsterdam*: Brain tissues were obtained from The Netherlands Brain Bank (NBB), Netherlands Institute for Neuroscience, Amsterdam (open access: www.brainbank.nl). All Material has been collected from donors for or from whom a written informed consent for a brain autopsy and the use of the material and clinical information for research purposes had been obtained by the NBB.

*DZNE:* Samples were provided from the Brain Bank Unit Tübingen of the DZNE Brain Bank. The DZNE Brain Bank is funded by the German Center of Neurodegenerative Diseases. JH is funded by the Deutsche Forschungsgemeinschaft (DFG, German Research Foundation) under Germany’s Excellence Strategy within the framework of the Munich Cluster for Systems Neurology (EXC 2145 SyNergy– ID 390857198).

*Stockholm*: The Brain bank at Karolinska Institutet receives CIMED-funding.

*Paris*: The NeuroCEB Neuropathology network includes: Drs Jean Boutonnat, Virginie Scolandre, & François Paysant (CHU Grenoble) ; Dr. Mathilde Duchesne (CHU Limoges) ; Dr Franck Letournel (CHU Angers), Pr Marie-Laure Martin-Négrier (CHU Bordeaux), Dr Maxime Faisant (CHU Caen), Pr Catherine Godfraind (CHU Clermont- Ferrand), Prs Claude-Alain Maurage & Vincent Deramecourt (CHU Lille), Pr David Meyronnet & Dr Romain Perbet (CHU Lyon) ; Dr Clémence Delteil (CHU Marseille), Dr Laurianne Geoffray (CHU Marseille) ; Pr Valérie Rigau (CHU Montpellier), Pr Danielle Seilhean, Drs Susana Boluda & Isabelle Plu (GH SU, Paris), Dr Dan Christian Chiforeanu (CHU Rennes), Dr Florent Marguet (CHU Rouen), Dr Béatrice Lannes (CHU Strasbourg). STL is funded by the National Institute on Aging (P30AG066546), Michael J. Fox Foundation (MJFF-020301), Texas Alzheimer’s Research and Care Consortium (2020-20-25-CR)

*Victoria*: Brain tissues were received from the Victorian Brain Bank, supported by The Florey, The Alfred, Victorian Institute of Forensic Medicine and Coroners Court of Victoria and funded in part by Parkinson’s Victoria, MND Victoria, FightMND, Yulgilbar Foundation and Ian and Maria Cootes.

*Sydney*: JBK is supported by NHMRC Dementia Team 1095127. GMH receives funding from NHMRC program grants 1037746 and 1132524, NHMRC Dementia Team 1095127, and NHMRC Fellowships 1079679 and 1176607. OP receives funding from by NHMRC program grant 1132524 and NHMRC Dementia Team 1095127, and NHMRC Fellowships 1103258 and 2008020. JJK and JRH both receive funding from NHMRC program grants 1037746 and 1132524, and NHMRC Dementia Team 1095127.

#### **(F) Bibliography**

1.	Boughton AP, Welch RP, Flickinger M, VandeHaar P, Taliun D, Abecasis GR, et al. LocusZoom.js: interactive and embeddable visualization of genetic association study results. Bioinformatics. 2021;37(18):3017–8.

2.	Real R, Martinez-Carrasco A, Reynolds RH, Lawton MA, Tan MMX, Shoai M, et al. Association between the LRP1B and APOE loci in the development of Parkinson’s disease dementia. Brain. 2023 May 2;146(5):1873-1887.

3.	Yang J, Lee SH, Goddard ME, Visscher PM. GCTA: A tool for genome-wide complex trait analysis. Am J Hum Genet. 2011;88(1):76–82.

4.	Zou Y, Carbonetto P, Wang G, Stephens M. Fine-mapping from summary data with the “Sum of Single Effects” model. PLoS Genet. 2022;18(7):1–24.

5.	Benner C, Spencer CCA, Havulinna AS, Salomaa V, Ripatti S, Pirinen M. FINEMAP: Efficient variable selection using summary data from genome-wide association studies. Bioinformatics. 2016;32(10):1493–501.

6.	de Klein N, Tsai EA, Vochteloo M, Baird D, Huang Y, Chen CY, et al. Brain expression quantitative trait locus and network analyses reveal downstream effects and putative drivers for brain-related diseases. Nat Genet. 2023 Mar;55(3):377-388.

7.	Giambartolomei C, Vukcevic D, Schadt EE, Franke L, Hingorani AD, Wallace C, et al. Bayesian Test for Colocalisation between Pairs of Genetic Association Studies Using Summary Statistics. PLoS Genet. 2014;10(5).

8.	Uhlén M, Fagerberg L, Hallström BM, Lindskog C, Oksvold P, Mardinoglu A, et al. Tissue-based map of the human proteome. Science (1979). 2015;347(6220).

9.	Berglund L, Björling E, Oksvold P, Fagerberg L, Asplund A, Szigyarto CAK, et al. A genecentric human protein atlas for expression profiles based on antibodies. Molecular and Cellular Proteomics. 2008;7(10):2019–27.

10.	Zhang Y, Sloan SA, Clarke LE, Caneda C, Plaza CA, Blumenthal PD, et al. Purification and Characterization of Progenitor and Mature Human Astrocytes Reveals Transcriptional and Functional Differences with Mouse. Neuron. 2016;89(1):37–53.

11.	Saunders A, Macosko EZ, Wysoker A, Goldman M, Krienen FM, de Rivera H, et al. Molecular Diversity and Specializations among the Cells of the Adult Mouse Brain. Cell. 2018;174(4):1015-1030.e16.

12.	Janer A, Martin E, Muriel MP, Latouche M, Fujigasaki H, Ruberg M, et al. PML clastosomes prevent nuclear accumulation of mutant ataxin-7 and other polyglutamine proteins. Journal of Cell Biology. 2006;174(1):65–76.

13.	Guo L. A cellular system that degrades misfolded proteins and protects against neurodegeneration. Mol Cell. 2014;

14.	Forlani G, Tosi G, Turrini F, Poli G, Vicenzi E, Accolla RS. Tripartite motif-containing protein 22 interacts with class II transactivator and orchestrates its recruitment in nuclear bodies containing TRIM19/PML and Cyclin T1. Front Immunol. 2017 May 15:8:564.

15.	Shaik S, Pandey H, Thirumalasetti SK, Nakamura N. Characteristics and Functions of the Yip1 Domain Family (YIPF), Multi-Span Transmembrane Proteins Mainly Localized to the Golgi Apparatus. Front Cell Dev Biol. 2019;7(July).

16.	Kuijpers M, Yu K Lou, Teuling E, Akhmanova A, Jaarsma D, Hoogenraad CC. The ALS8 protein VAPB interacts with the ER-Golgi recycling protein YIF1A and regulates membrane delivery into dendrites. EMBO Journal. 2013 Jul 17;32(14):2056–72.

17.	AlMuhaizea M, AlMass R, AlHargan A, AlBader A, Medico Salsench E, Howaidi J, et al. Truncating mutations in YIF1B cause a progressive encephalopathy with various degrees of mixed movement disorder, microcephaly, and epilepsy. Acta Neuropathol. 2020;139(4):791–4.

18.	Medico Salsench E, Maroofian R, Deng R, Lanko K, Nikoncuk A, Pérez B, et al. Expanding the mutational landscape and clinical phenotype of the YIF1B related brain disorder. Brain. 2021;144(10).

19.	Sergeant N, Sablonnière B, Schraen-Maschke S, Ghestem A, Maurage CA, Wattez A, et al. Dysregulation of human brain microtubule-associated tau mRNA maturation in myotonic dystrophy type 1. Hum Mol Genet. 2001;10(19):2143–55.

20.	Caillet-Boudin ML, Fernandez-Gomez FJ, Tran H, Dhaenens CM, Buee L, Sergeant N. Brain pathology in myotonic dystrophy: When tauopathy meets spliceopathy and RNAopathy. Front Mol Neurosci. 2014 Jan 9:6:57.

21.	Sabry S, Vuillaumier-Barrot S, Mintet E, Fasseu M, Valayannopoulos V, Héron D, et al. A case of fatal Type I congenital disorders of glycosylation (CDG I) associated with low dehydrodolichol diphosphate synthase (DHDDS) activity. Orphanet J Rare Dis. 2016 Jun 24;11(1):84.

22.	Mehta S, Lal V. DHDDS Mutation: A Rare Cause of Refractory Epilepsy and Hyperkinetic Movement Disorder. J Mov Disord. 2023 Jan;16(1):107-109.

23.	Galosi S, Edani BH, Martinelli S, Hansikova H, Eklund EA, Caputi C, et al. De novo DHDDS variants cause a neurodevelopmental and neurodegenerative disorder with myoclonus. Brain. 2022 Mar 29;145(1):208-223.
